## Supplementary Materials for "Characterizing the interactions between influenza and respiratory syncytial viruses and their implications for epidemic control"

### Supplementary Figures

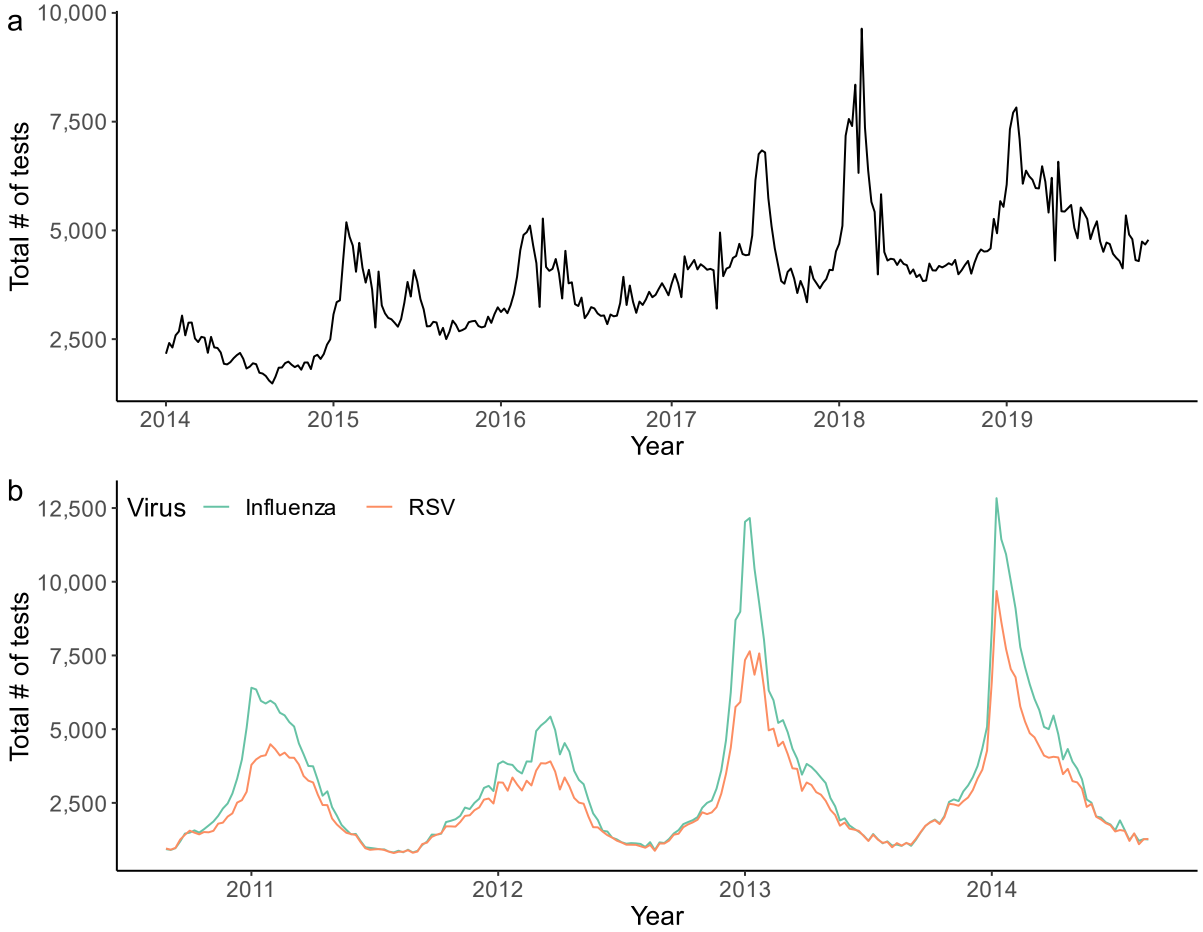


**Supplementary Figure 1. Weekly number of tests performed for influenza and RSV in Hong Kong (a) and Canada (b) throughout the study period.** In Hong Kong, all samples were tested for both viruses. In Canada, samples tested for influenza are shown in green, and samples tested for RSV are shown in orange.

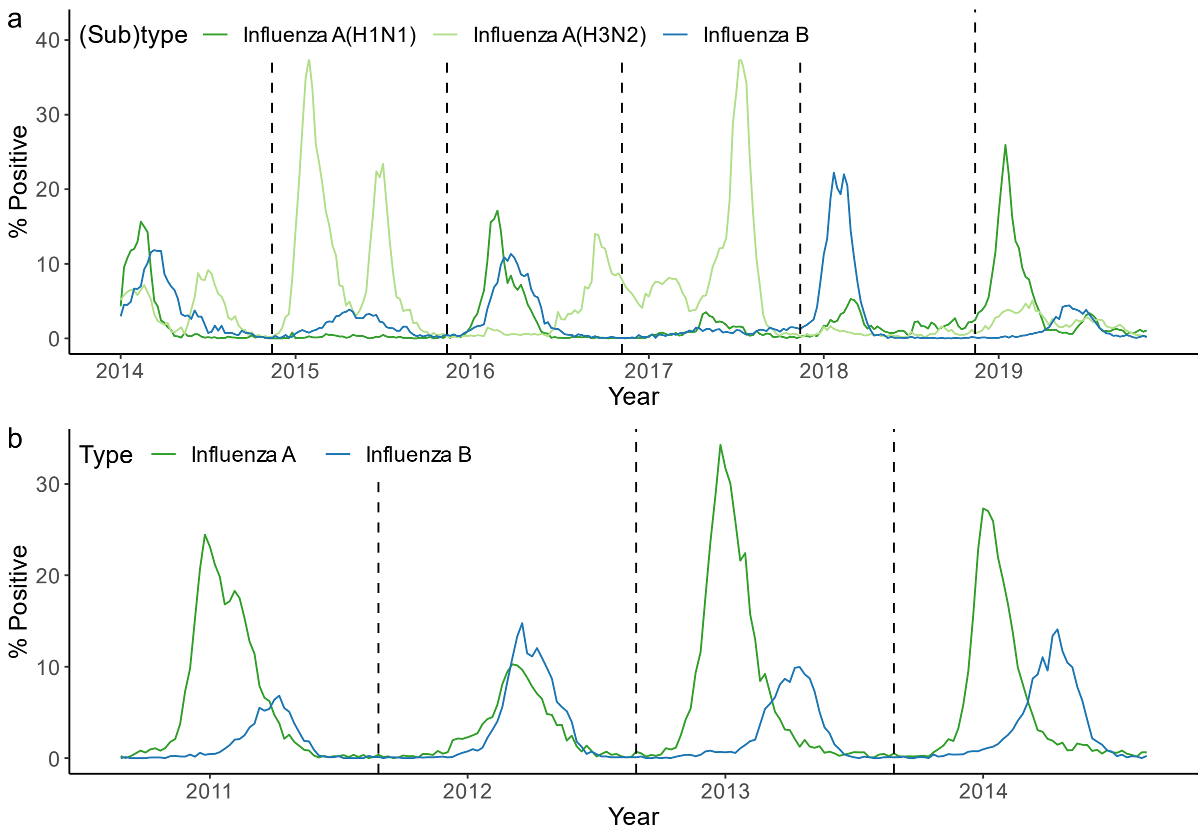


**Supplementary Figure 2. Percent of tests positive for influenza by week and (sub)type in Hong Kong (a) and Canada (b) throughout the study period.** Values for influenza A are shown in green and values for influenza B are shown in blue. For Hong Kong, A(H1N1) data are shown in dark green and A(H3N2) data in light green; in Canada, subtyping of influenza A samples was not consistently conducted.

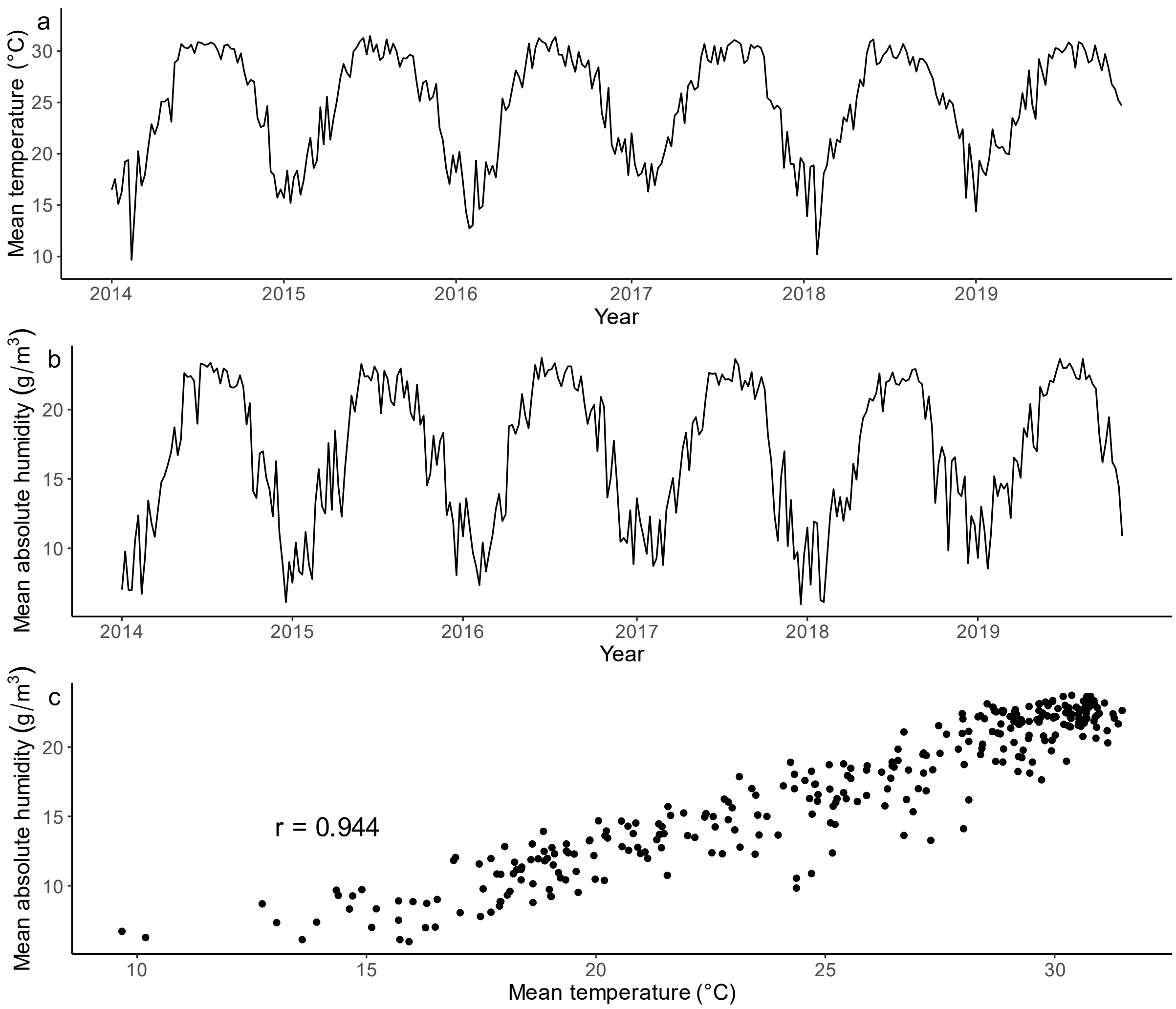


**Supplementary Figure 3. Weekly temperature and absolute humidity in Hong Kong.** This figure shows the weekly average (a) temperature (in degrees Celsius) and (b) absolute humidity (in grams per meter cubed) in Hong Kong throughout the study period. The relationship between temperature and absolute humidity, as well as the corresponding Pearson correlation coefficient, is shown in (c).

**Supplementary Figure 4. Maximum likelihood estimates of season-specific parameters in Hong Kong (a) and Canada (b).** Points represent the maximum likelihood estimates, while error bars represent the 95% confidence intervals, as obtained through parametric bootstrapping using 500 synthetic datasets. Note that, on occasion, the MLEs fall outside of the confidence intervals (points shown in red), although only slightly. The meaning of each season-specific parameter is described in Table 1 in the main text; $R_{10} + R_{120}$ refers to the total proportion of the population immune to influenza at the beginning of the season, while $R_{20} + R_{120}$ is the same but for RSV.

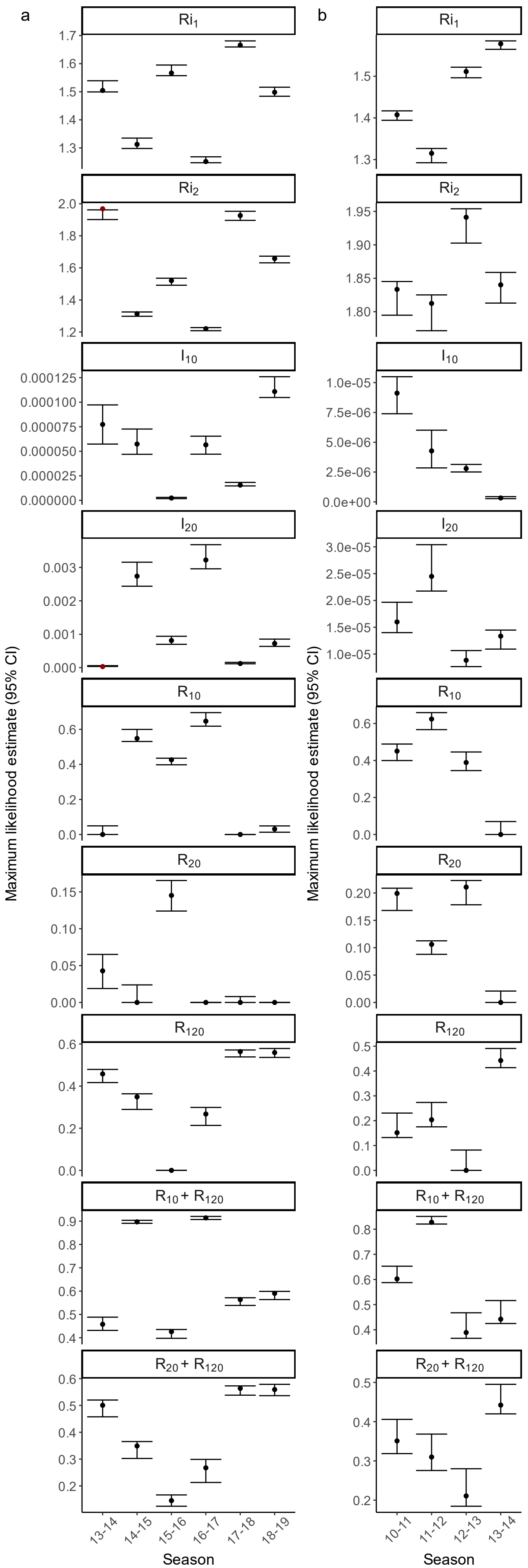

a
b
Ri1
Ri2
I10
I20
R10
R20
R120
R10 + R120
R20 + R120
Maximum likelihood estimate (95% CI)
Season
13-14
14-15
15-16
16-17
17-18
18-19
10-11
11-12
12-13

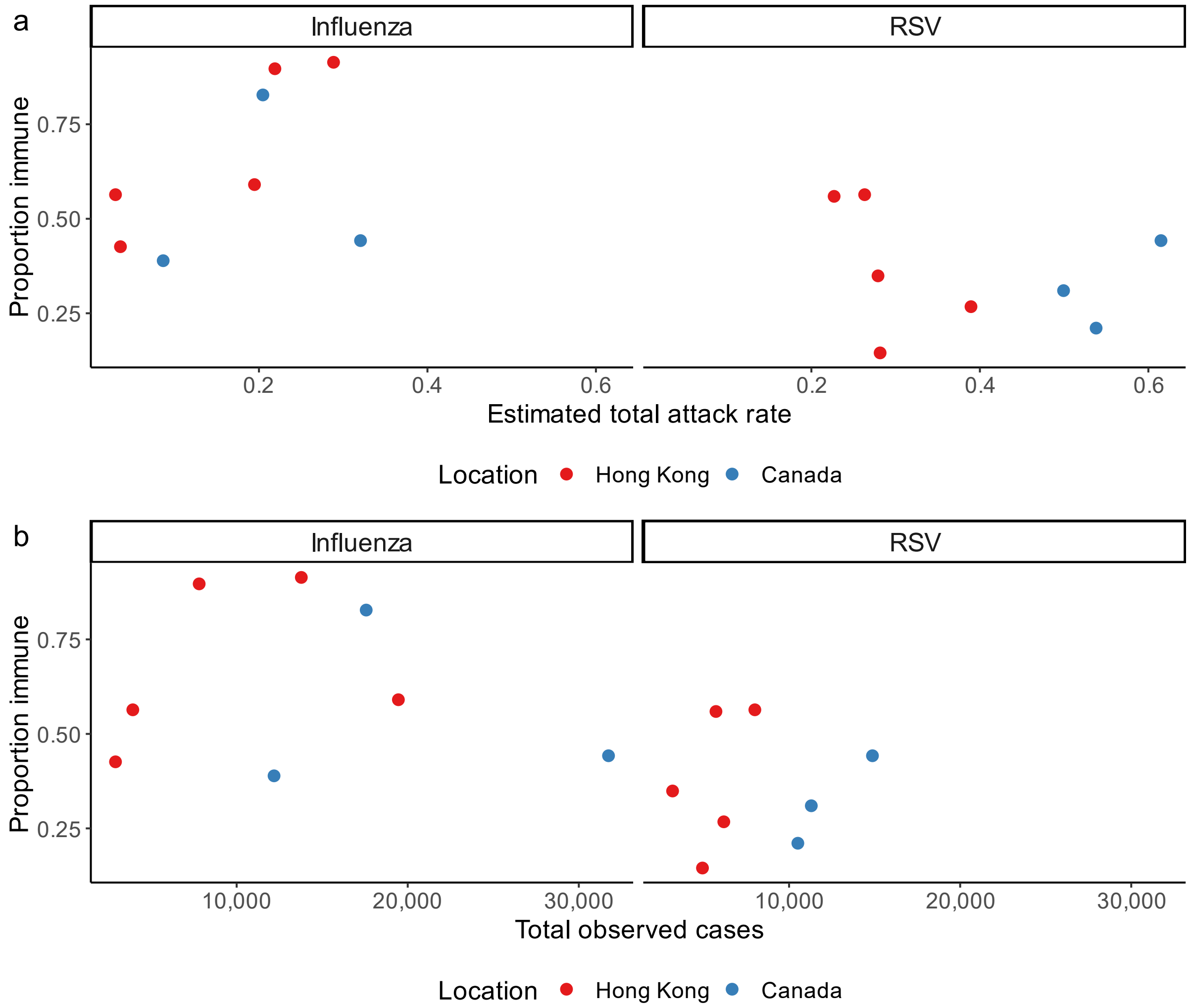


**Supplementary Figure 5. Relationship between the previous season's attack rate and the proportion immune to influenza and RSV at the beginning of each season.** The relationship with the total proportion immune is shown for both (a) the estimated total attack rate, as calculated by running the deterministic model at the maximum likelihood estimate for all seasons, and (b) the observed number of cases in the raw data. Values from Hong Kong are shown in red, while values from Canada are shown in blue. For each location, the earliest season (2013-14 for Hong Kong and 2010-11 for Canada) is not included, as no data were available prior to these seasons.

**Supplementary Figure 6. Correlations between best-fit values of all shared model parameters.** Results from Hong Kong are shown in (a), and results from Canada in (b).Each point in the lower left panels represents the fit values of shared model parameters from a single fit of the model. Correlation coefficients (Kendall's $\tau$) between the fit values of each pair of shared parameters are shown in the upper right panels; significance is indicated using dots and asterisks (. $p<0.10$; * $p<0.05$; ** $p<0.01$; *** $p<0.001$). Density plots of the top fits for each parameter are shown on the diagonal. All fits falling within the 95% confidence interval (defined based on a chi-squared distribution with 54 (Hong Kong) or 40 (Canada) degrees of freedom, according to Wilks' theorem) are included.

a

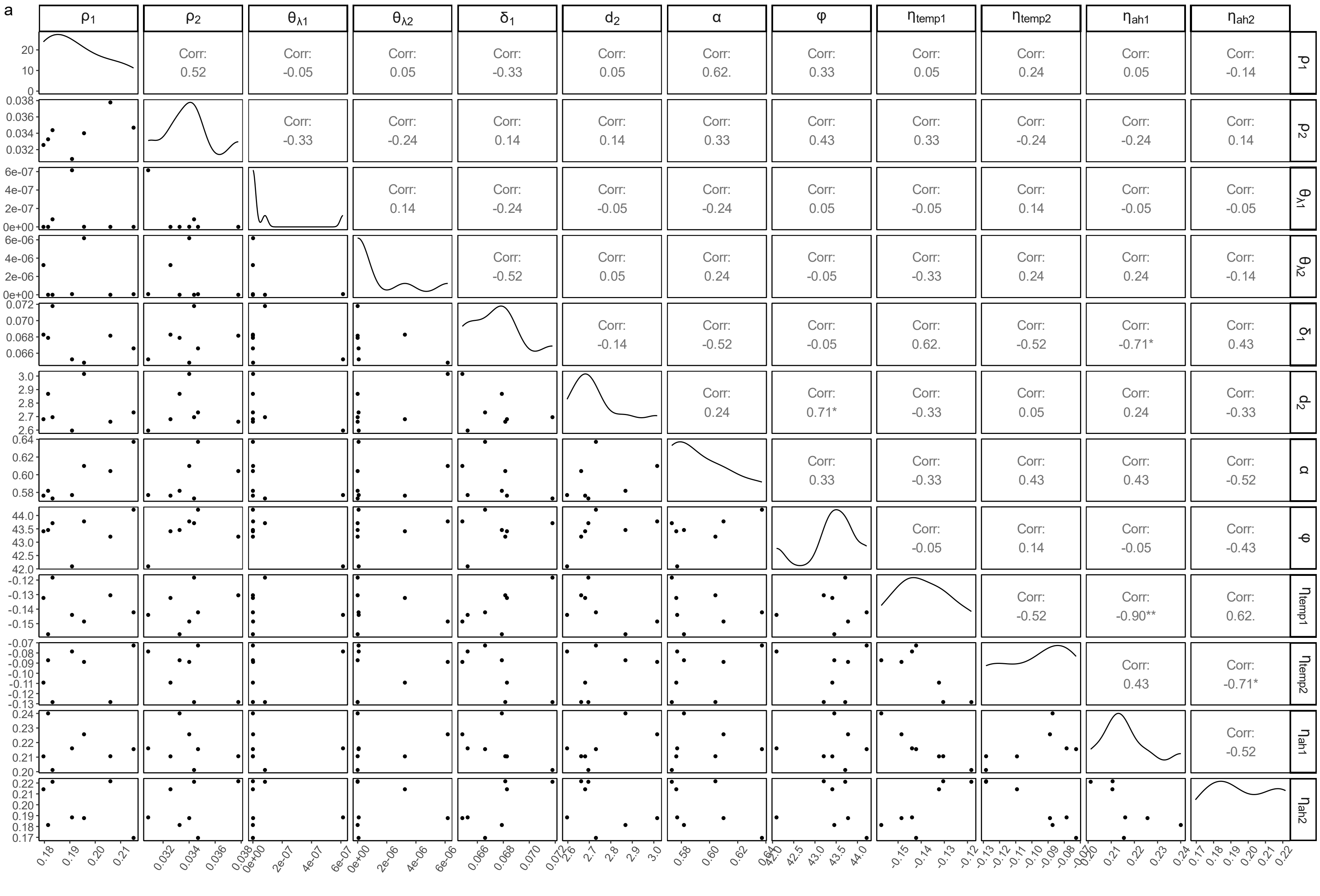

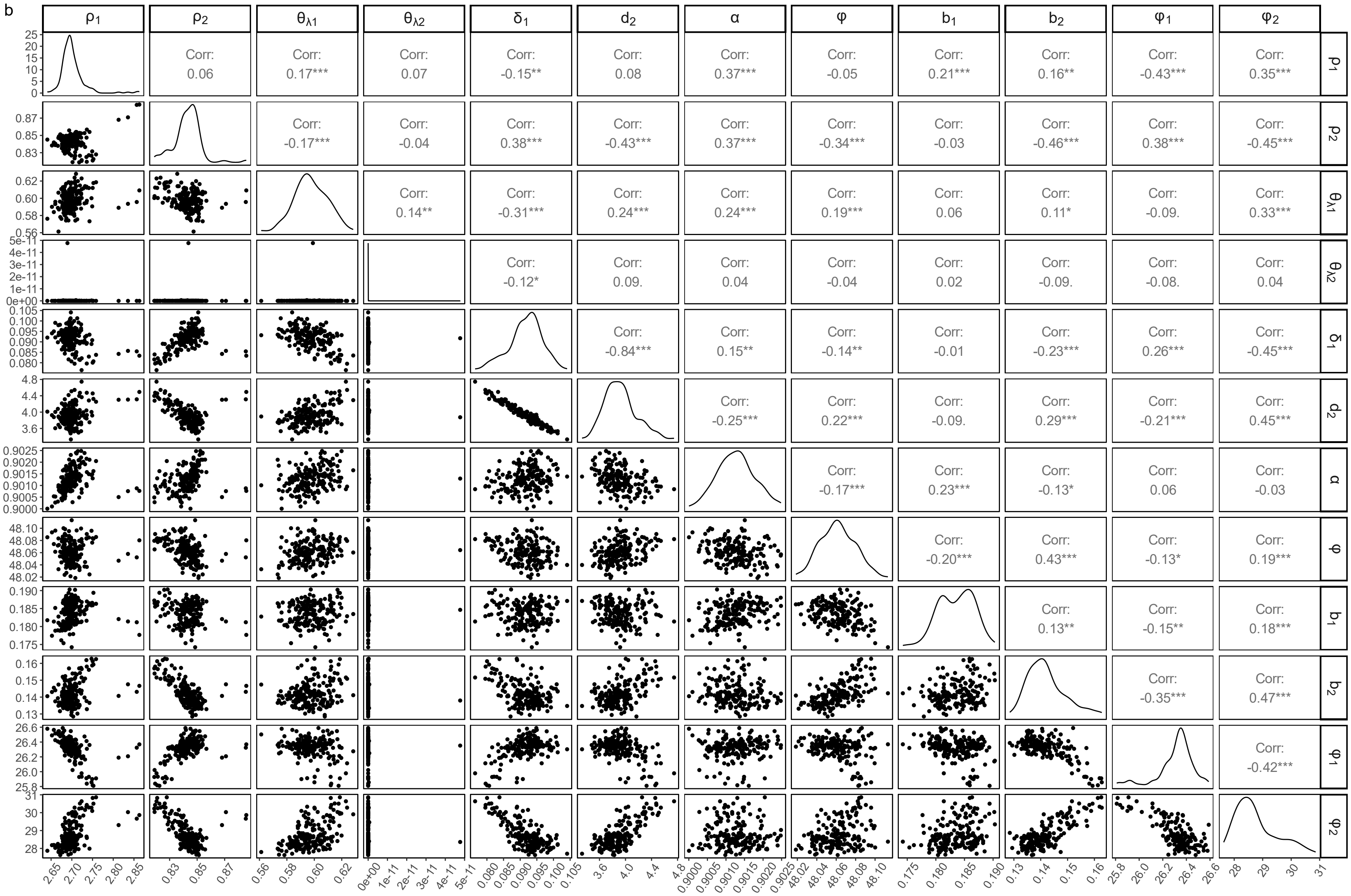

b
ρ1
ρ2
θλ1
θλ2
δ1
d2
α
φ
b1
b2
φ1
φ2
Corr: 0.06
Corr: 0.17***
Corr: 0.07
Corr: -0.15**
Corr: 0.08
Corr: 0.37***
Corr: -0.05
Corr: 0.21***
Corr: 0.16**
Corr: -0.43***
Corr: 0.35***
Corr: -0.17***
Corr: -0.04
Corr: 0.38***
Corr: -0.43***
Corr: 0.37***
Corr: -0.34***
Corr: -0.03
Corr: -0.46***
Corr: 0.38***
Corr: -0.45***
Corr: 0.14**
Corr: -0.31***
Corr: 0.24***
Corr: 0.24***
Corr: 0.19***
Corr: 0.06
Corr: 0.11*
Corr: -0.09.
Corr: 0.33***
Corr: -0.12*
Corr: 0.09.
Corr: 0.04
Corr: -0.04
Corr: 0.02
Corr: -0.09.
Corr: -0.08.
Corr: 0.04
Corr: -0.84***
Corr: 0.15**
Corr: -0.14**
Corr: -0.01
Corr: -0.23***
Corr: 0.26***
Corr: -0.45***
Corr: -0.25***
Corr: 0.22***
Corr: -0.09.
Corr: 0.29***
Corr: -0.21***
Corr: 0.45***
Corr: -0.17***
Corr: 0.23***
Corr: -0.13*
Corr: 0.06
Corr: -0.03
Corr: -0.20***
Corr: 0.43***
Corr: -0.13*
Corr: 0.19***
Corr: 0.13**
Corr: -0.15**
Corr: 0.18***
Corr: -0.35***
Corr: 0.47***
Corr: -0.42***

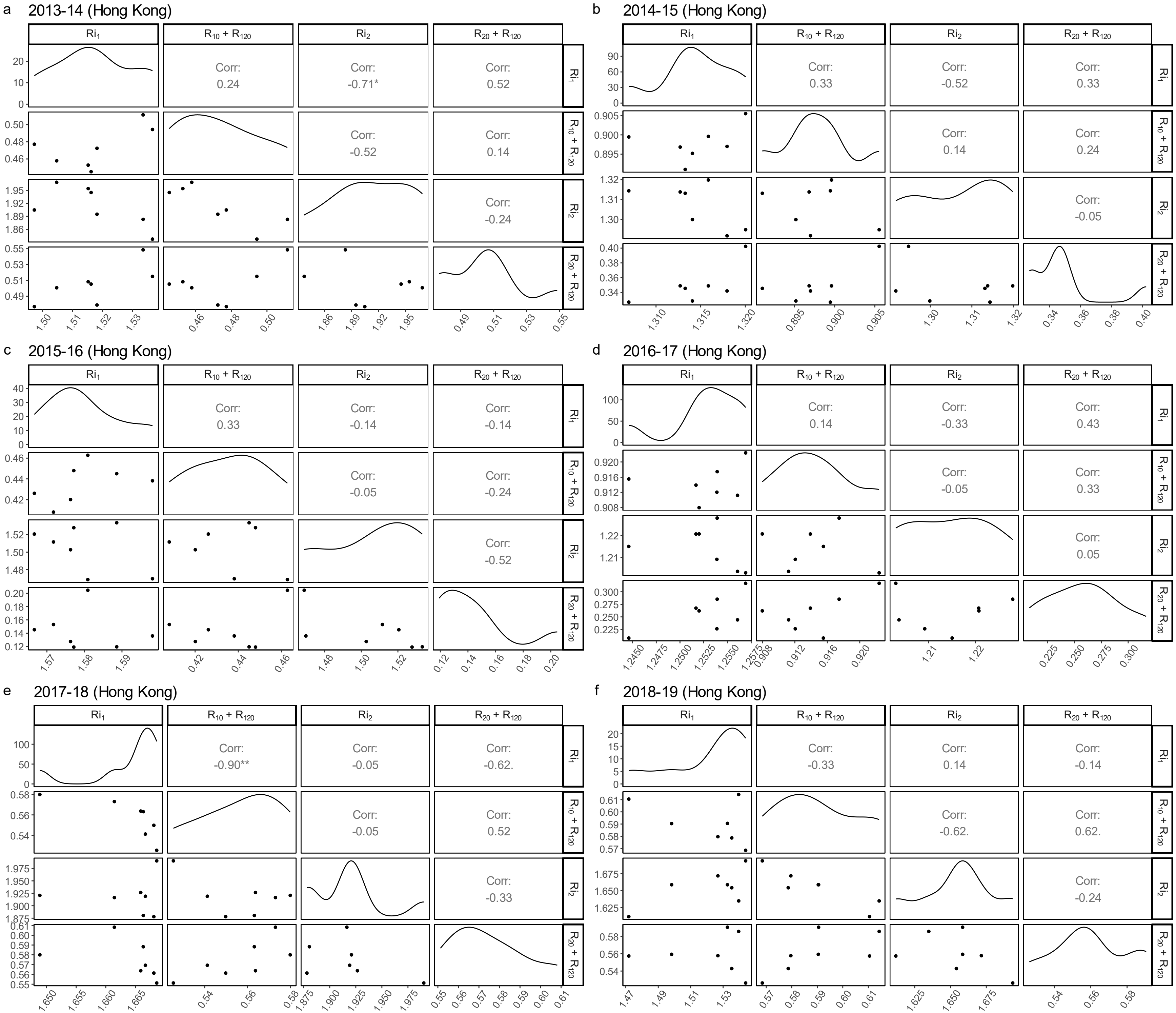
a
2013-14 (Hong Kong)
Ri$_1$
R$_{10}$ + R$_{120}$
Ri$_2$
R$_{20}$ + R$_{120}$
Corr: 0.24
Corr: -0.71*
Corr: 0.52
Corr: -0.52
Corr: 0.14
Corr: -0.24
b
2014-15 (Hong Kong)
Ri$_1$
R$_{10}$ + R$_{120}$
Ri$_2$
R$_{20}$ + R$_{120}$
Corr: 0.33
Corr: -0.52
Corr: 0.33
Corr: 0.14
Corr: 0.24
Corr: -0.05
c
2015-16 (Hong Kong)
Ri$_1$
R$_{10}$ + R$_{120}$
Ri$_2$
R$_{20}$ + R$_{120}$
Corr: 0.33
Corr: -0.14
Corr: -0.14
Corr: -0.05
Corr: -0.24
Corr: -0.52
d
2016-17 (Hong Kong)
Ri$_1$
R$_{10}$ + R$_{120}$
Ri$_2$
R$_{20}$ + R$_{120}$
Corr: 0.14
Corr: -0.33
Corr: 0.43
Corr: -0.05
Corr: 0.33
Corr: 0.05
e
2017-18 (Hong Kong)
Ri$_1$
R$_{10}$ + R$_{120}$
Ri$_2$
R$_{20}$ + R$_{120}$
Corr: -0.90**
Corr: -0.05
Corr: -0.62.
Corr: -0.05
Corr: 0.52
Corr: -0.33
f
2018-19 (Hong Kong)
Ri$_1$
R$_{10}$ + R$_{120}$
Ri$_2$
R$_{20}$ + R$_{120}$
Corr: -0.33
Corr: 0.14
Corr: -0.14
Corr: -0.62.
Corr: 0.62.
Corr: -0.24

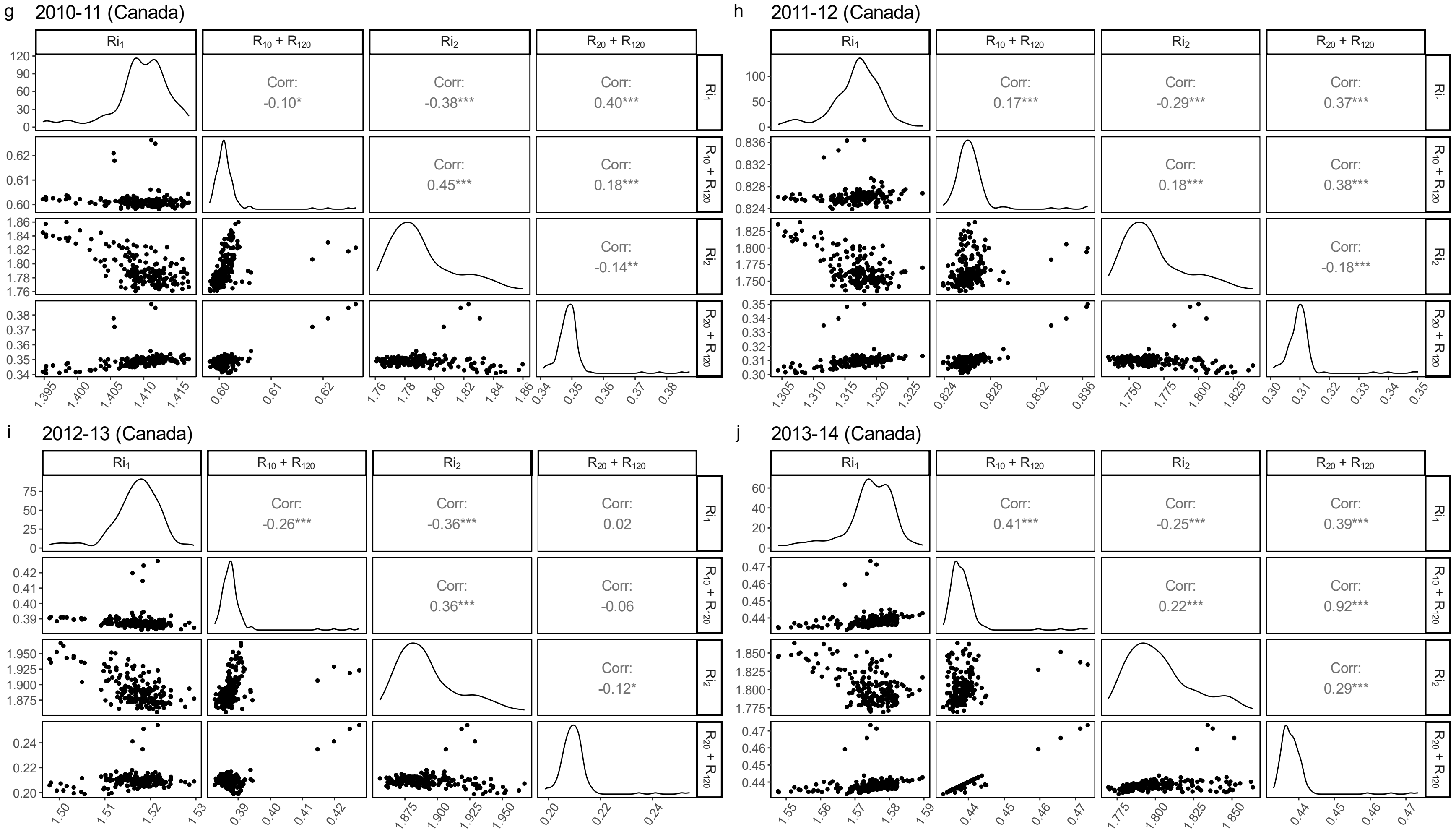
g 2010-11 (Canada)
Ri1
R10 + R120
Ri2
R20 + R120
Corr: -0.10*
Corr: -0.38***
Corr: 0.40***
Corr: 0.45***
Corr: 0.18***
Corr: -0.14**
h 2011-12 (Canada)
Ri1
R10 + R120
Ri2
R20 + R120
Corr: 0.17***
Corr: -0.29***
Corr: 0.37***
Corr: 0.18***
Corr: 0.38***
Corr: -0.18***
i 2012-13 (Canada)
Ri1
R10 + R120
Ri2
R20 + R120
Corr: -0.26***
Corr: -0.36***
Corr: 0.02
Corr: 0.36***
Corr: -0.06
Corr: -0.12*
j 2013-14 (Canada)
Ri1
R10 + R120
Ri2
R20 + R120
Corr: 0.41***
Corr: -0.25***
Corr: 0.39***
Corr: 0.22***
Corr: 0.92***
Corr: 0.29***

**Supplementary Figure 7. Correlations between best-fit values of season-specific model parameters.** Each panel (a-j) displays results for an individual season in either Hong Kong (a-f) or Canada (g-j). Similarly to Supplementary Fig. 6, points in the lower left panels represent the fit values of the season-specific model parameters from a single model fit, while correlation coefficients (Kendall's τ) between the fit values of each parameter pair are shown in the upper right panels. Significance is indicated using dots and asterisks (. p<0.10; * p<0.05; ** p<0.01; *** p<0.001). Density plots of the top fits for each parameter are shown on the diagonal. Again, all fits falling within the 95% confidence interval are included.

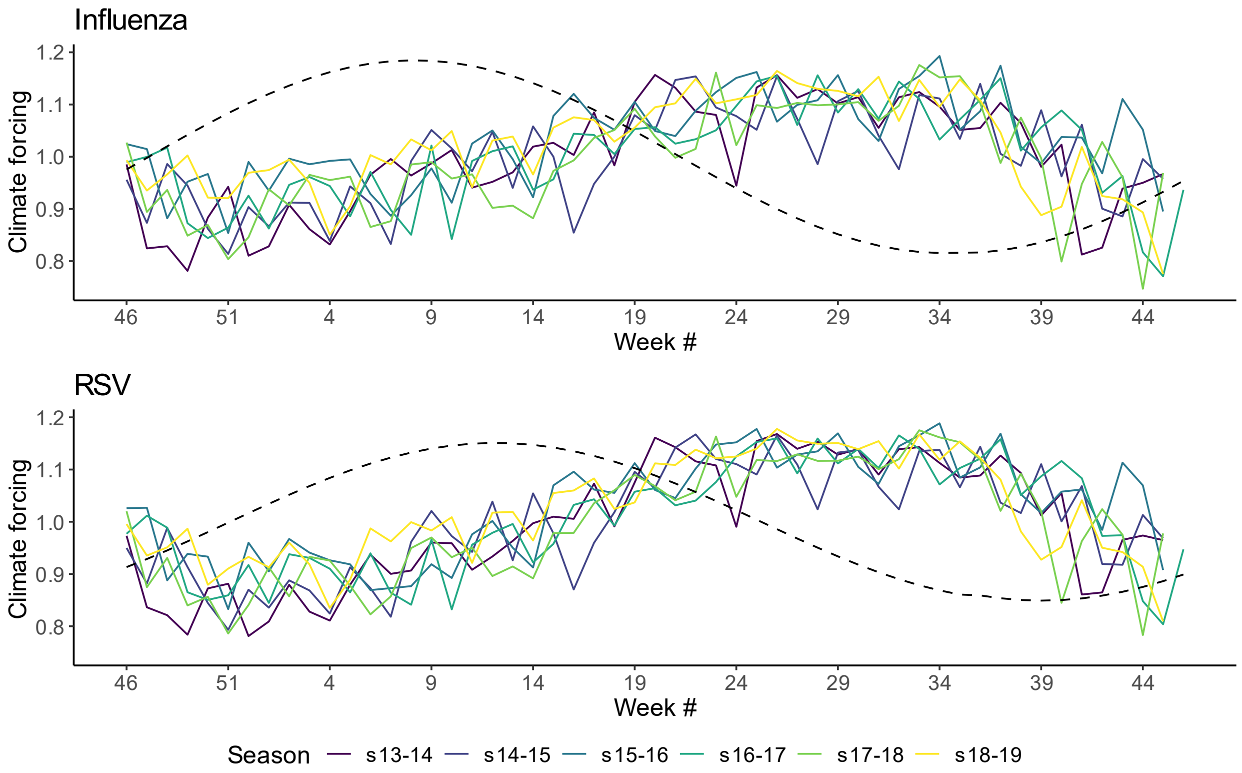


**Supplementary Figure 8. Extent of climate forcing of influenza and RSV transmission by week and season.** Values for each week and season were calculated using the maximum likelihood values (see Table 1) of $\eta_{temp1}$ and $\eta_{AH1}$ for influenza, and of $\eta_{temp2}$ and $\eta_{AH2}$ for RSV, as well as the standardized temperature and absolute humidity data from Hong Kong. In the model, these values are multiplied by $\widehat{\beta}_i$ to obtain the weekly force of infection (see Equation (1) in the main text). Line colors indicate season. The extent of sinusoidal forcing fit in Canada is shown as dotted lines, for comparison.

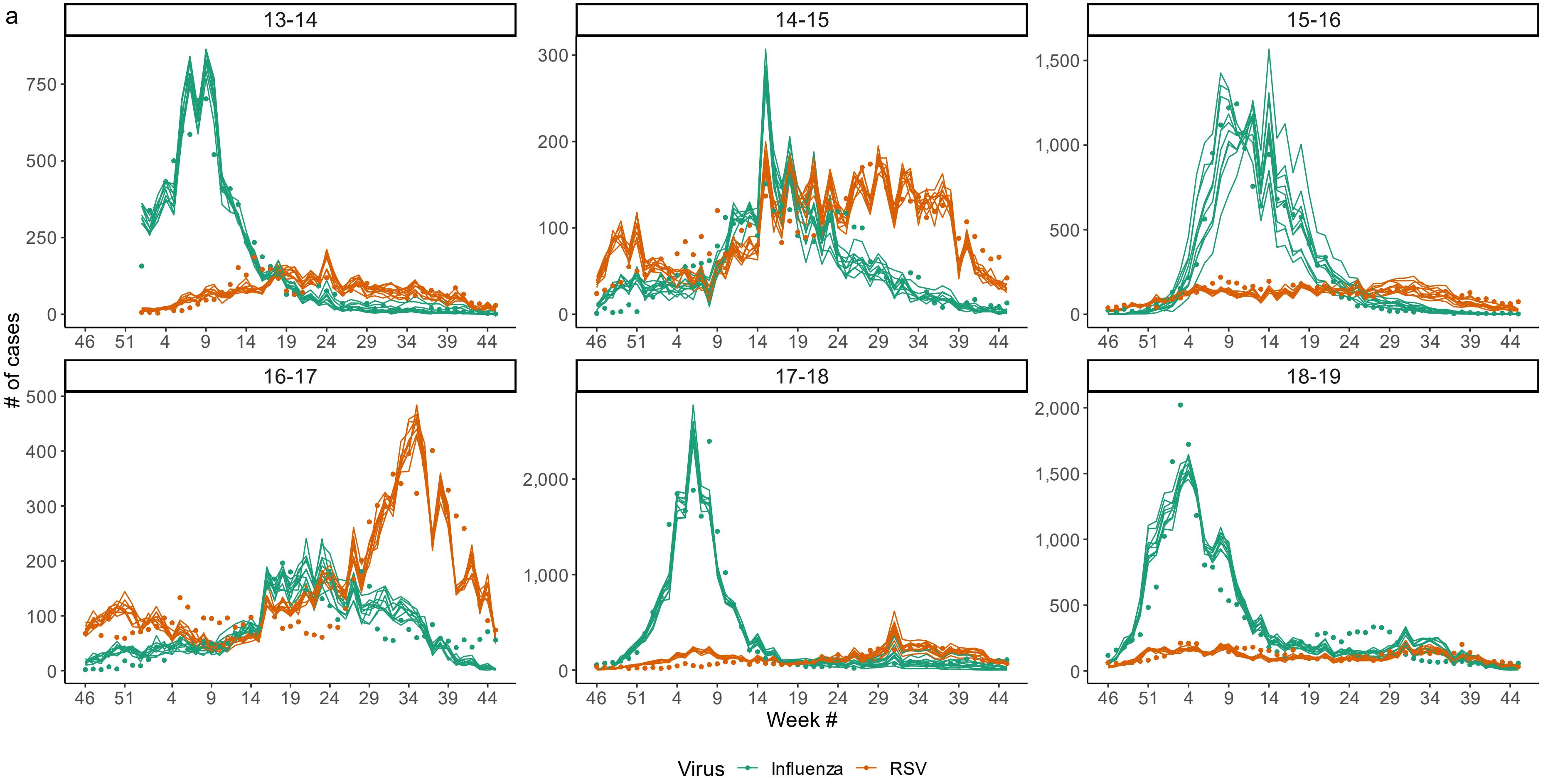

a
13-14
14-15
15-16
16-17
17-18
18-19
### of cases
Week #
Virus
Influenza
RSV

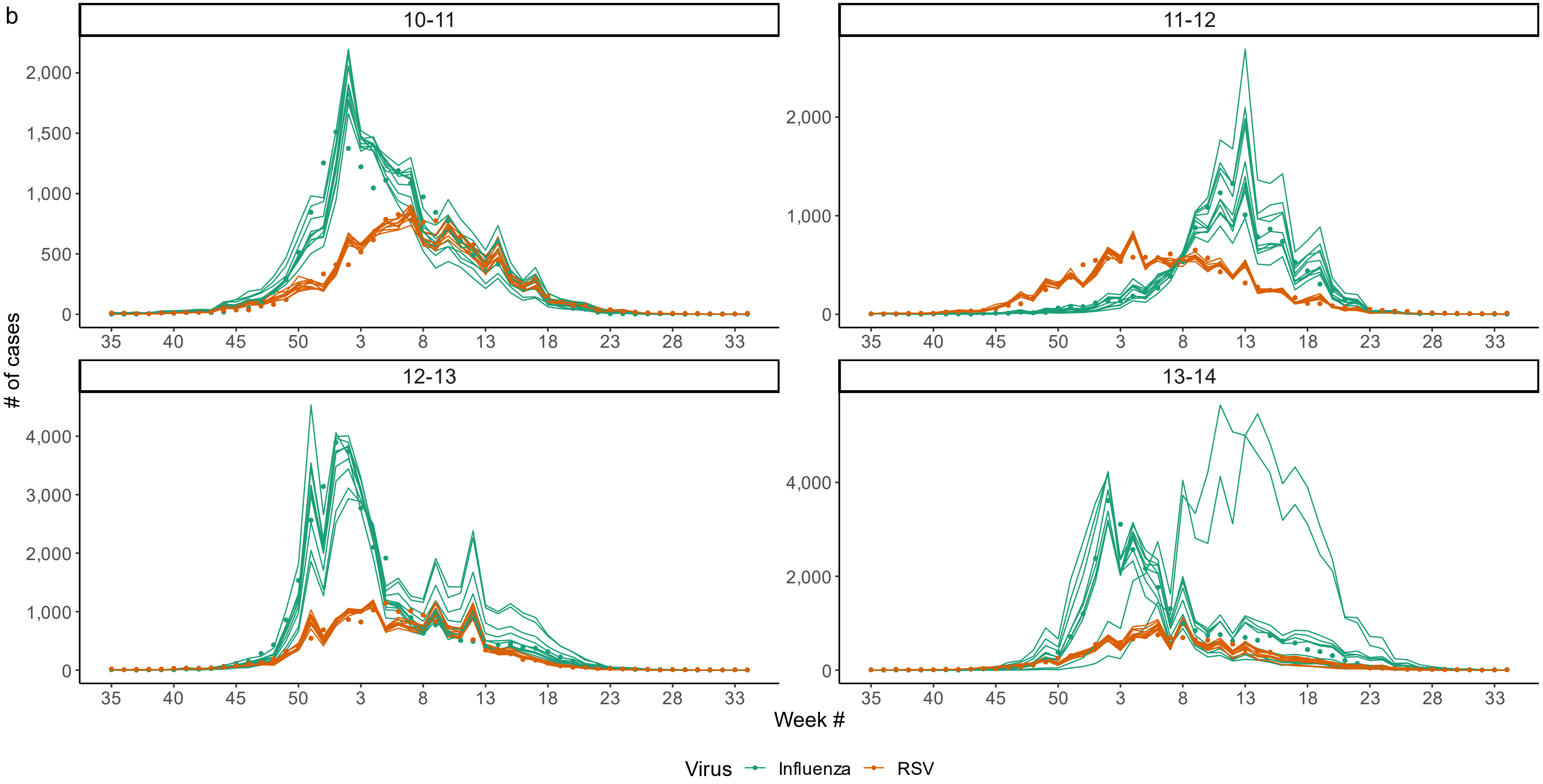


**Supplementary Figure 9. Model simulations at the maximum likelihood estimate in Hong Kong (a) and Canada (b).** Simulations were run after setting all parameters to their maximum likelihood estimates (Table 1). Observed case counts are plotted as points, while ten realizations from the stochastic transmission model for each location and season are shown as lines. Colors chosen for each virus are as in Fig. 1.

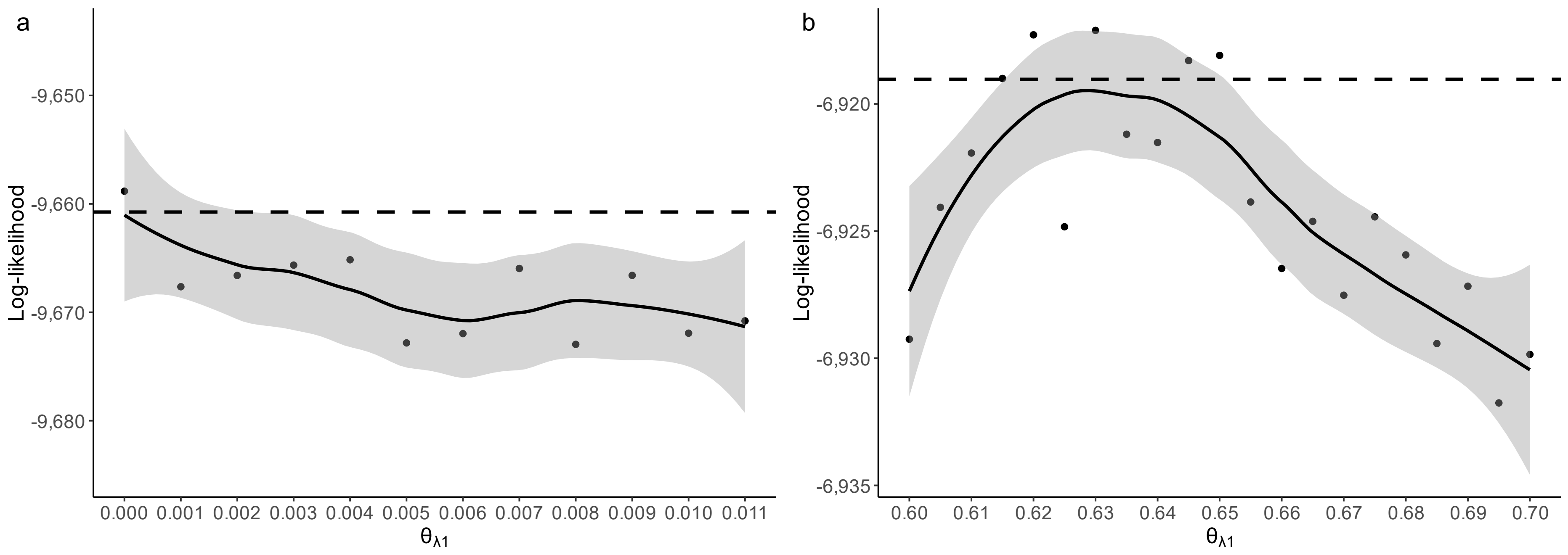


**Supplementary Figure 10. Profile likelihoods of $\theta_{\lambda 1}$ in Hong Kong (a) and Canada (b).** Log-likelihood values of the best model fits achieved when $\theta_{\lambda 1}$ was held constant at values between 0 and 0.11 (a) and between 0.6 and 0.7 (b) are shown as points. In (a), parameter values above 0.11 consistently had log-likelihoods much lower than the maximum values (i.e., a difference of >20) and were therefore not plotted. The solid lines represent smooths calculated using locally estimated scatterplot smoothing (LOESS) to fit the points, and the shaded areas represent the 95% confidence intervals of these smooths. Dotted horizontal lines represent the cutoff value beneath which fits no longer fall within the 95% confidence interval of the $\theta_{\lambda 1}$ value providing the best-quality fit, as defined based on a chi-square distribution with one degree of freedom.

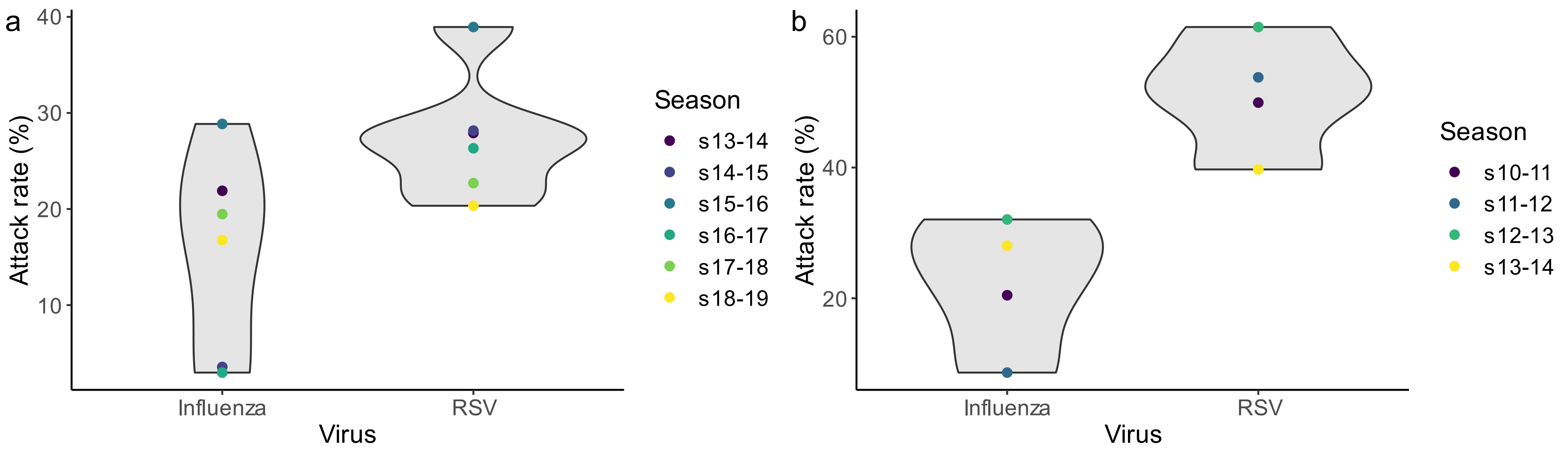


**Supplementary Figure 11. Simulated total attack rates of influenza and RSV at the maximum likelihood estimates in Hong Kong (a) and Canada (b).** Violin plots represent the range of estimated influenza and RSV attack rates, obtained by simulating the deterministic model with parameters set to their maximum likelihood values (Table 1), across all seasons for a given location. Values for individual seasons are shown as points.

**Supplementary Figure 12. Epidemic trajectories in the presence and absence of an interaction between influenza and RSV.** Deterministic simulations of RSV (a, c) and influenza (b, d) epidemics in the presence (darker lines) and absence (lighter lines) of the inferred interaction effect are shown for Hong Kong (a-b) and Canada (c-d). Simulations in the presence of the interaction were run using the maximum likelihood estimates (Table 1) of all model parameters; simulations in the absence of the interaction effect were conducted by setting the initial number of cases of the interacting pathogen (influenza in (a, c) and RSV in (b, d)) to 0, such that no outbreak occurred. In (a) and (c), influenza epidemics are displayed using dotted lines; in (b) and (d), dotted lines show the RSV epidemics.

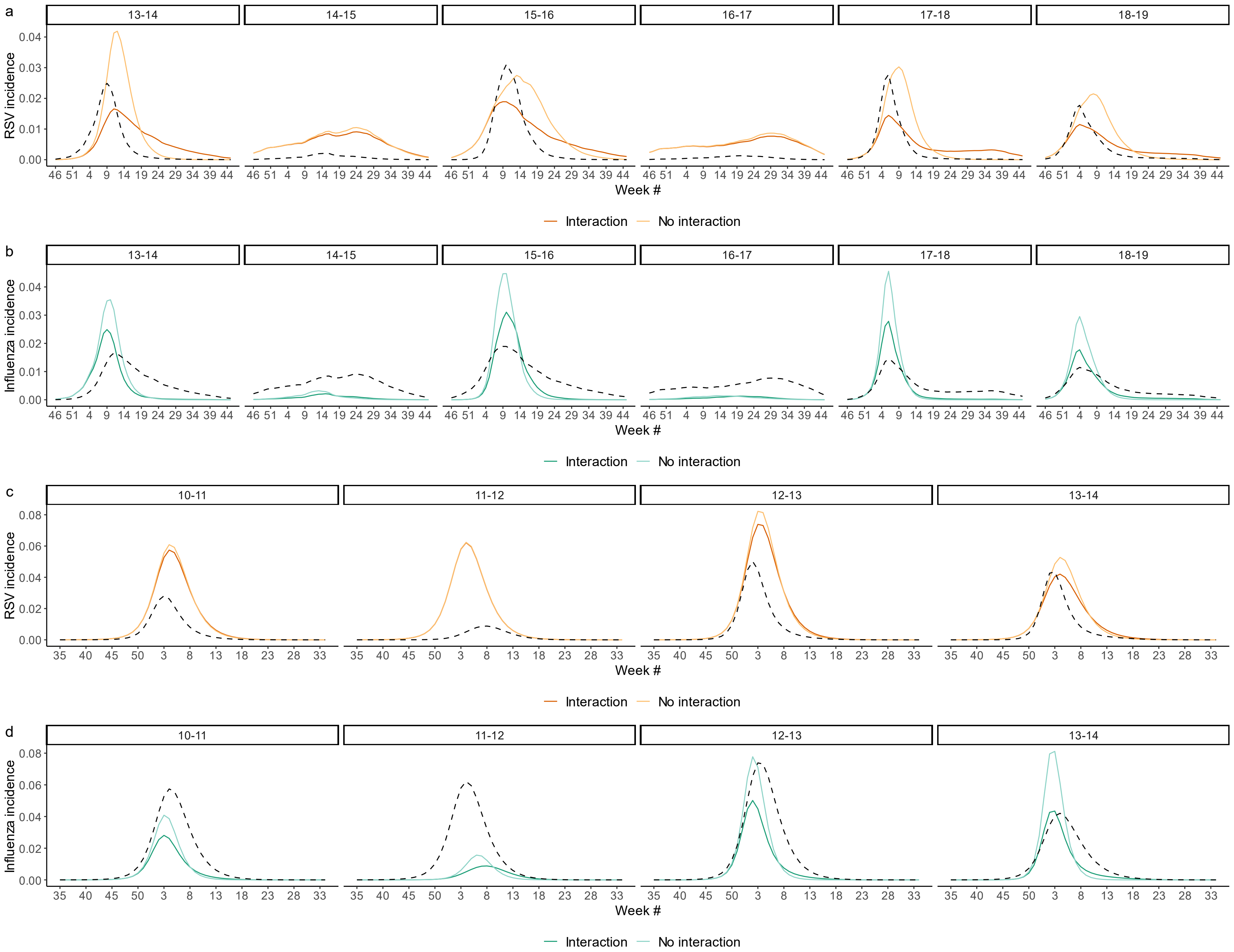
a
13-14
14-15
15-16
16-17
17-18
18-19
RSV incidence
0.04
0.03
0.02
0.01
0.00
46 51 4 9 14 19 24 29 34 39 44
Week #
Interaction
No interaction
b
Influenza incidence
c
10-11
11-12
12-13
13-14
0.08
0.06
35 40 45 50 3 8 13 18 23 28 33
d

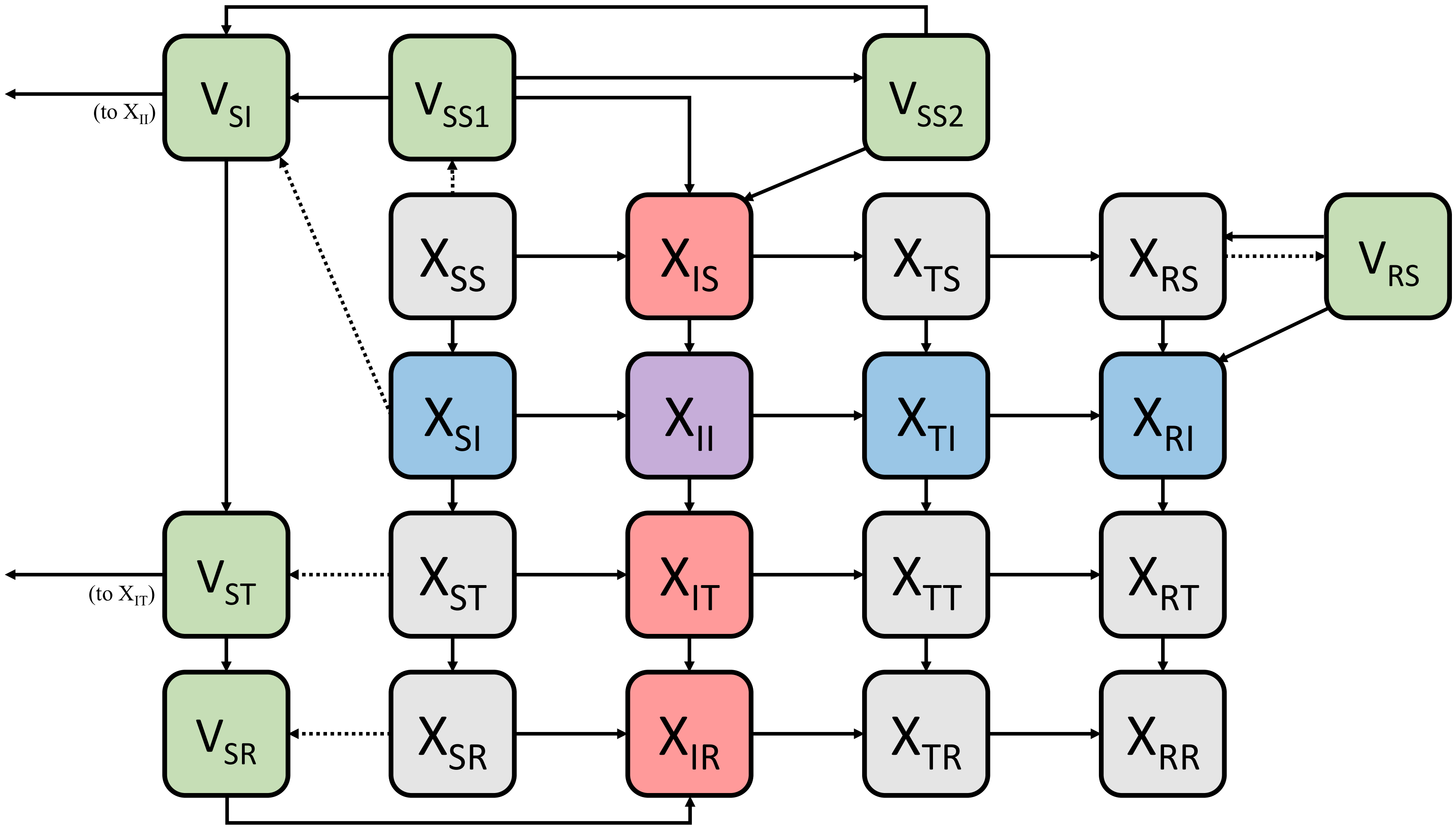


**Supplementary Figure 13. Model schematic for the simulation study of LAIV impact.** As in Fig. 2a, boxes represent model states, and arrows represent possible transitions between compartments. Dotted arrows represent the transitions undergone upon vaccination; these transitions occur instantaneously at a single time point, and have no further bearing on the transmission dynamics. Compartments containing vaccinated individuals are shown in green and labeled "V," whereas compartments with unvaccinated individuals are labeled "X." Subscripts indicate infection status with regard to influenza and RSV. Red shading indicates infection with influenza, blue shading indicates infection with RSV, and purple shading indicates coinfection.

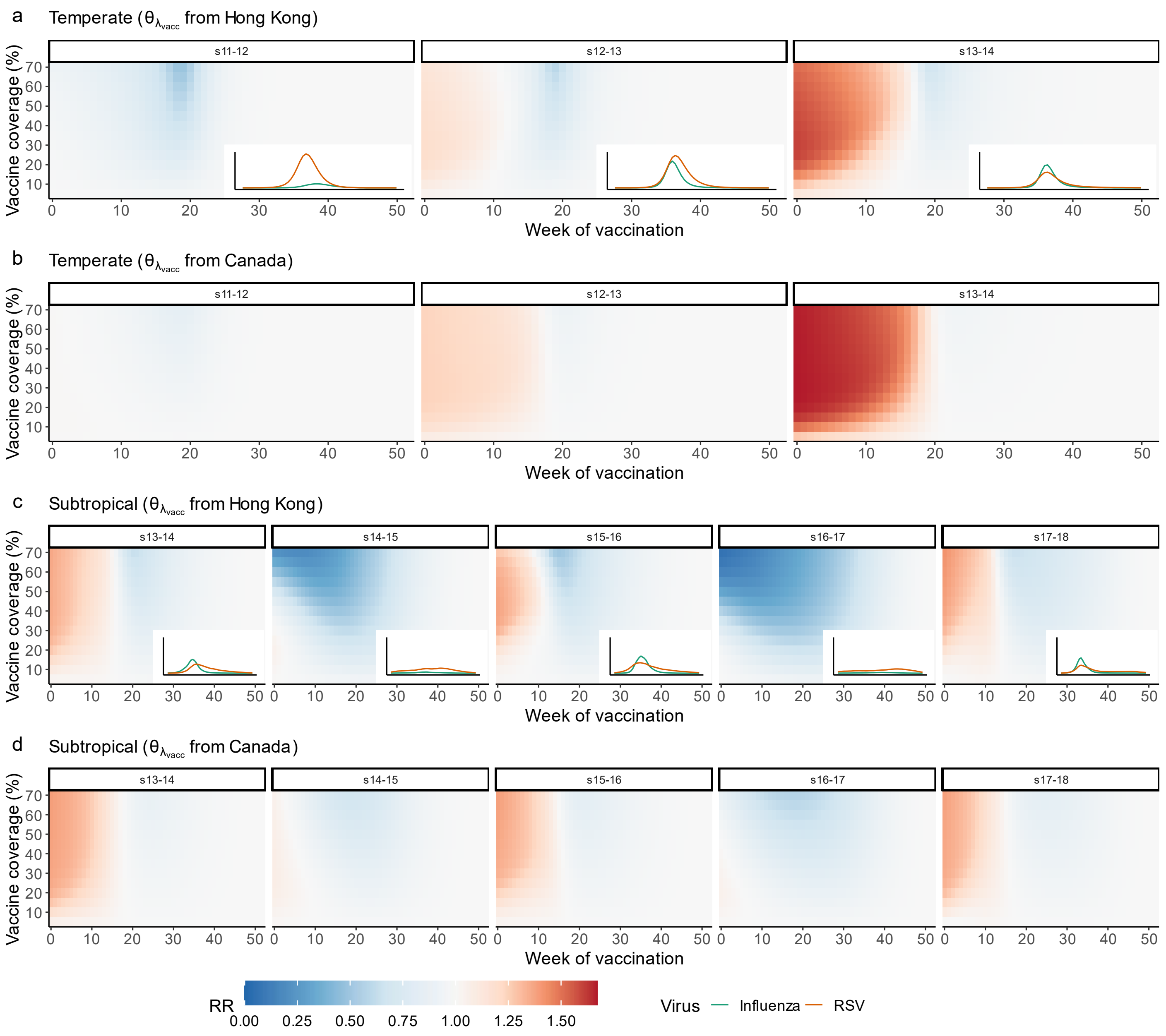

a
Temperate ($\theta_{\lambda_{vacc}}$ from Hong Kong)
b
Temperate ($\theta_{\lambda_{vacc}}$ from Canada)
c
Subtropical ($\theta_{\lambda_{vacc}}$ from Hong Kong)
d
Subtropical ($\theta_{\lambda_{vacc}}$ from Canada)
s11-12
s12-13
s13-14
s14-15
s15-16
s16-17
s17-18
Vaccine coverage (%)
Week of vaccination
RR
0.00
0.25
0.50
0.75
1.00
1.25
1.50
Virus
Influenza
RSV

**Supplementary Figure 14. Impact of LAIV on RSV attack rates by season.** Heatmaps show the impact of LAIV on RSV attack rates for all seasons not included in Fig. 4 for both the temperate (a-b) and subtropical (c-d) scenarios at various values of vaccine timing and coverage. Vaccination was assumed to either strongly reduce RSV susceptibility (a, c), as found in Hong Kong, or to moderately reduce RSV susceptibility (b, d), as found in Canada. Values below 1.0 are indicated in blue, while values above 1.0 are indicated in red. Inset plots in A and C show the weekly percentage of the model population infected with influenza (green) and RSV (orange) in the absence of vaccination for a given scenario and season. The maximum value for all inset plots is 6.2%, to allow comparison of outbreak magnitude across plots.

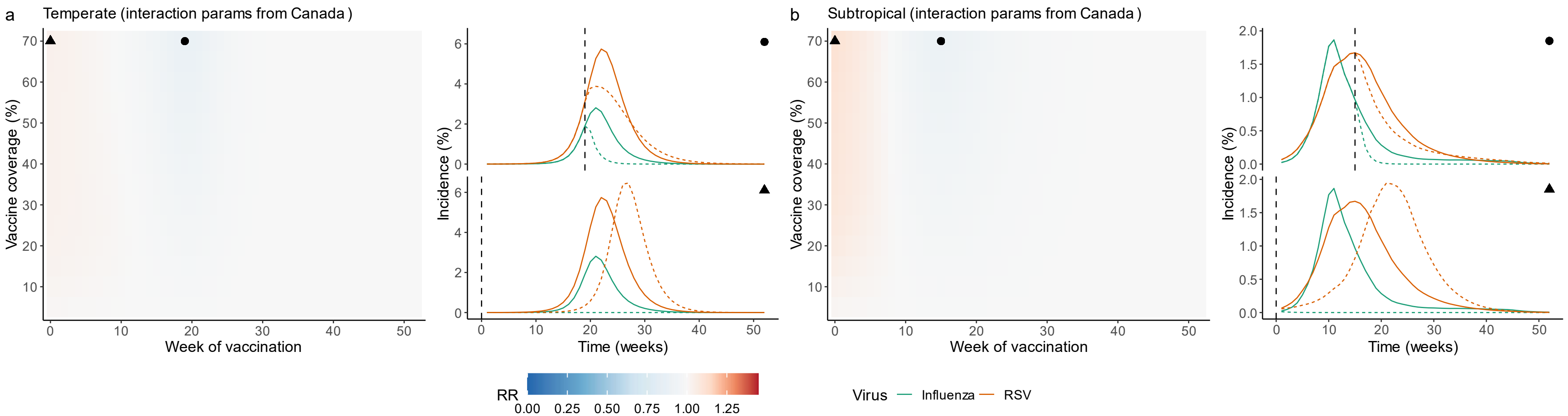


**Supplementary Figure 15. Impact of LAIV on RSV attack rates for the temperate (a) and subtropical (b) scenarios, assuming interaction parameters are as fit in Canada.** As in Fig. 4, heatmaps show the impact of LAIV on RSV attack rates, lines indicate the simulated incidence of influenza (green) and RSV (orange) in the absence (solid lines) and presence (dotted lines) of vaccination, and circles and triangles indicate the vaccine timing and coverage levels that led to the greatest reduction and the greatest increase in RSV burden, respectively. In the heatmaps, blue indicates a reduction in RSV attack rates, and red indicates an increase. The strength and duration of the vaccine effect on RSV are assumed to be the same as the impact of natural influenza infection, as fit in Canada.

**Supplementary Figure 16. Impact of LAIV on RSV attack rates as duration of protection and vaccine efficacy against influenza vary.** Heatmaps indicate the impact of LAIV on RSV attack rates, where blue boxes indicate a reduction in attack rates, and red boxes indicate an increase. (a, b) Results for a vaccine leading to a reduction in RSV susceptibility lasting 1 month. (c, d) Results for a vaccine leading to a reduction in RSV susceptibility lasting 6 months. (e, f) Results for a vaccine with 60% efficacy against influenza infection. (g, h) Results for a vaccine with 95% efficacy against influenza infection. For all panels, the impact of vaccination on susceptibility to RSV is set to be equal to the impact of natural influenza infection, as inferred in Hong Kong. As in Fig. 4, results for the temperate scenario (a, c, e, g) are taken from the 2010-11 season, while results for the subtropical scenario (b, d, f, h) are taken from the 2018-19 season.

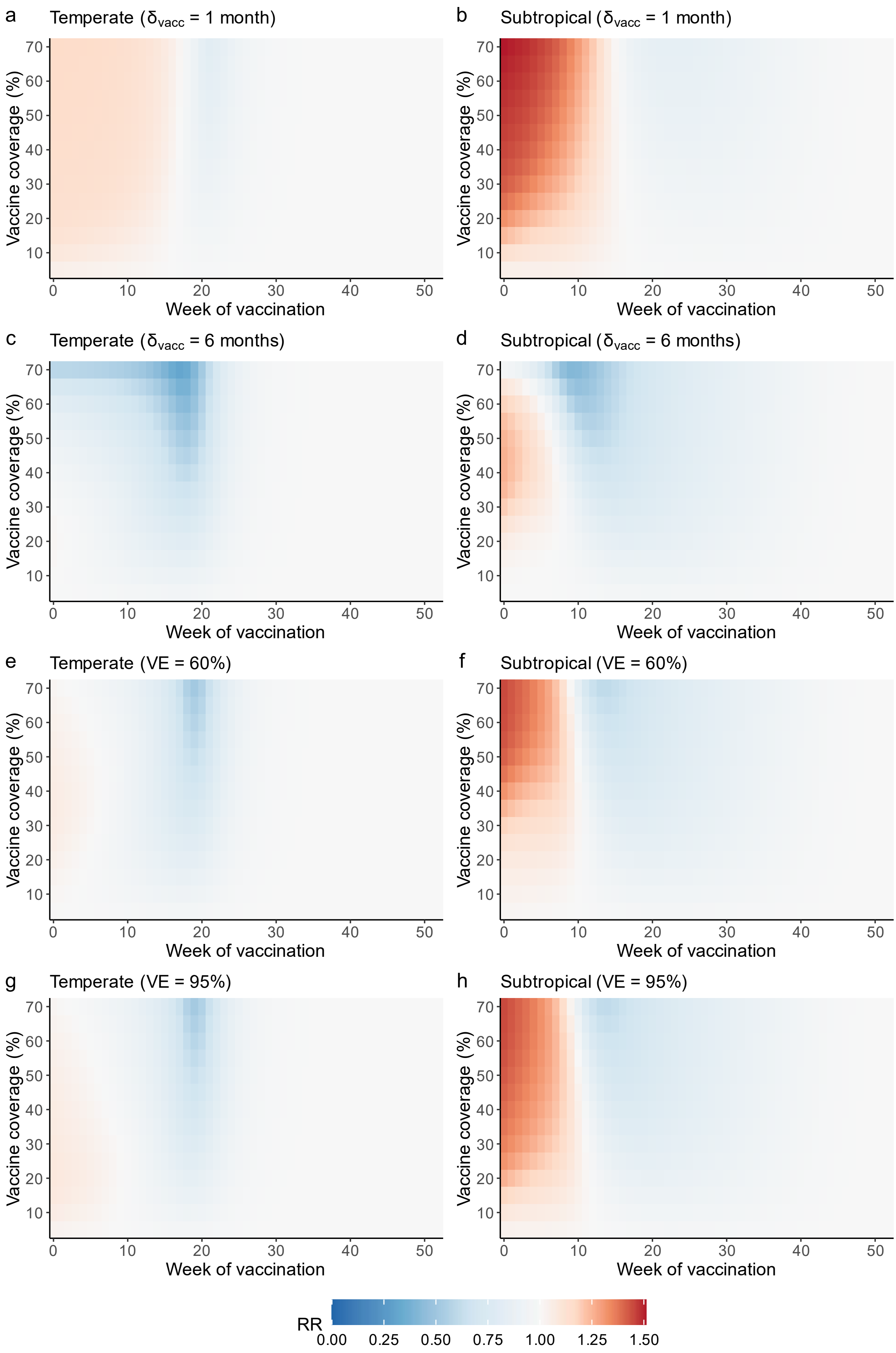

a
Temperate ($\delta_{vacc}$ = 1 month)
b
Subtropical ($\delta_{vacc}$ = 1 month)
c
Temperate ($\delta_{vacc}$ = 6 months)
d
Subtropical ($\delta_{vacc}$ = 6 months)
e
Temperate (VE = 60%)
f
Subtropical (VE = 60%)
g
Temperate (VE = 95%)
h
Subtropical (VE = 95%)
Vaccine coverage (%)
10
20
30
40
50
60
70
Week of vaccination
0
10
20
30
40
50
RR
0.00
0.25
0.50
0.75
1.00
1.25
1.50

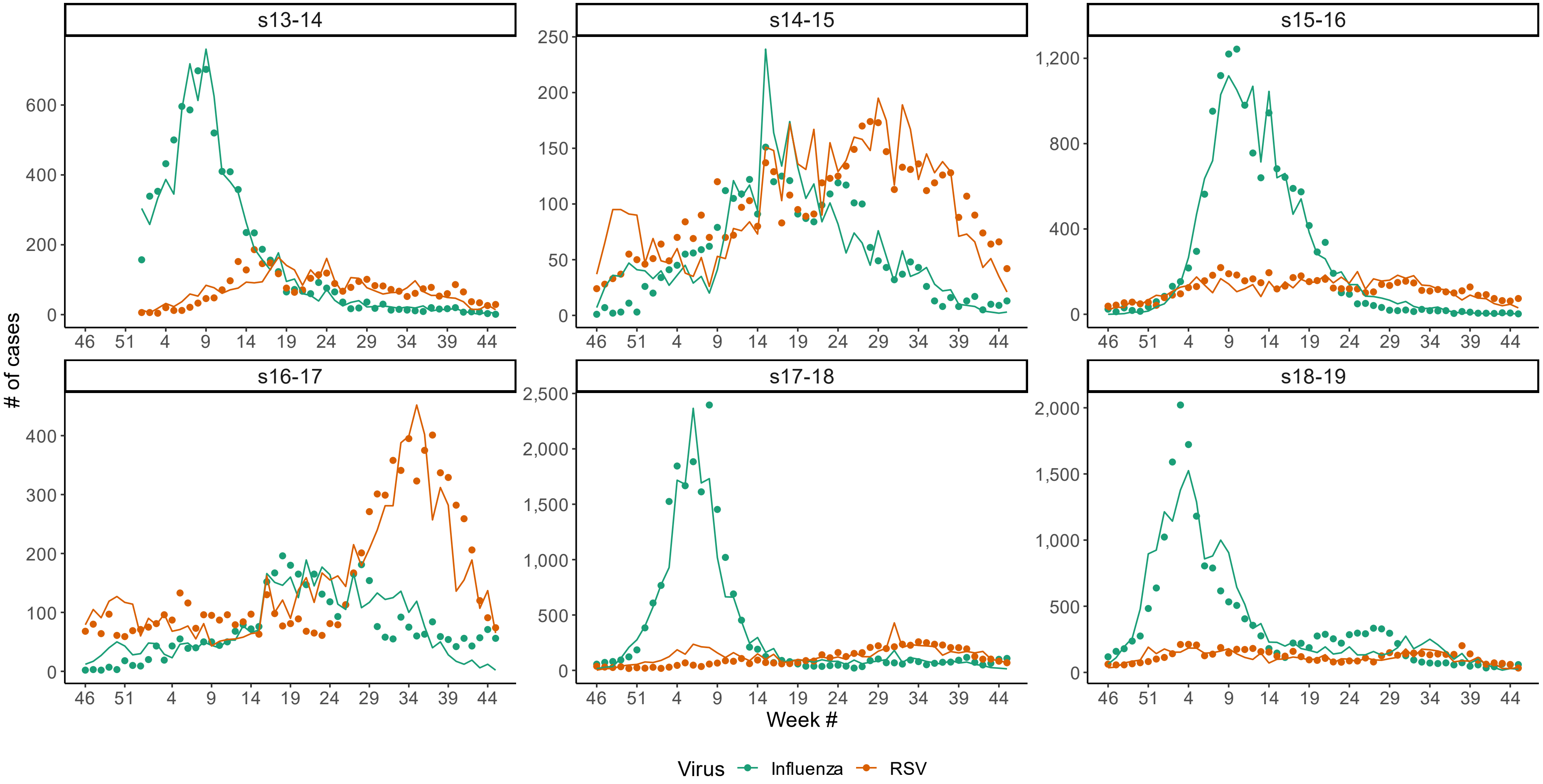


**Supplementary Figure 17. Comparison between observed data and synthetic data generated from the age-structured model.** For each season, lines represent the synthetic data generated from the age-structured model at the MLE and summed over all age groups, while points represent the total observed number of cases reported in the data. Synthetic data were generated by first running the deterministic age-structured model to obtain the total number of influenza and RSV cases in each age group, then drawing stochastically each week from a binomial distribution, with the number of trials equal to the total number of tests estimated to be conducted in a given age group that week, and the probability equal to $P_i(t)$ (main text Equations (3) and (4)) calculated for that age group and week.

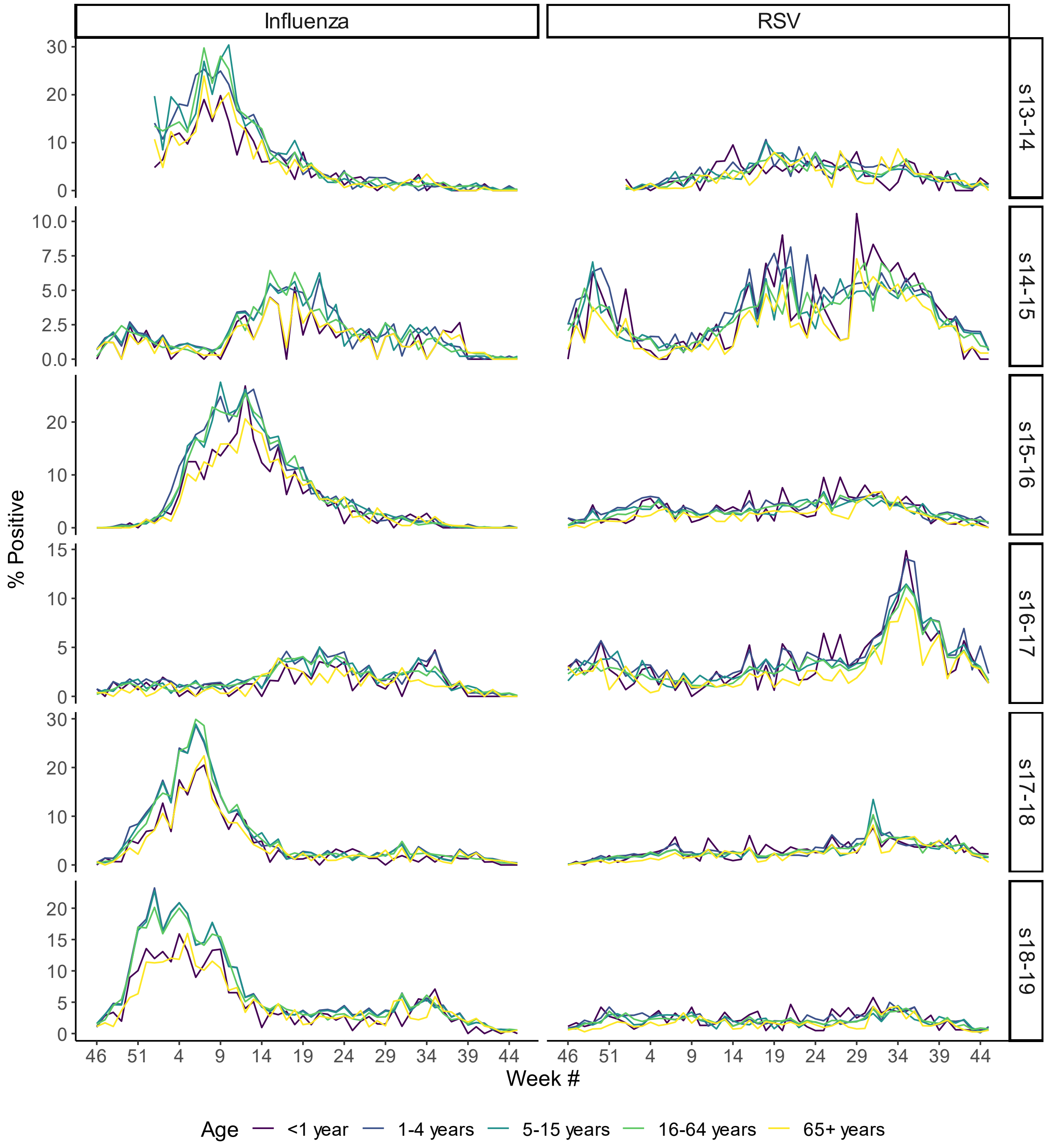


**Supplementary Figure 18. Synthetic data generated by the age-structured model.** The lines show the weekly percentage of tests positive for influenza (left) and RSV (right) for each season, by age. Line colors indicate age group.

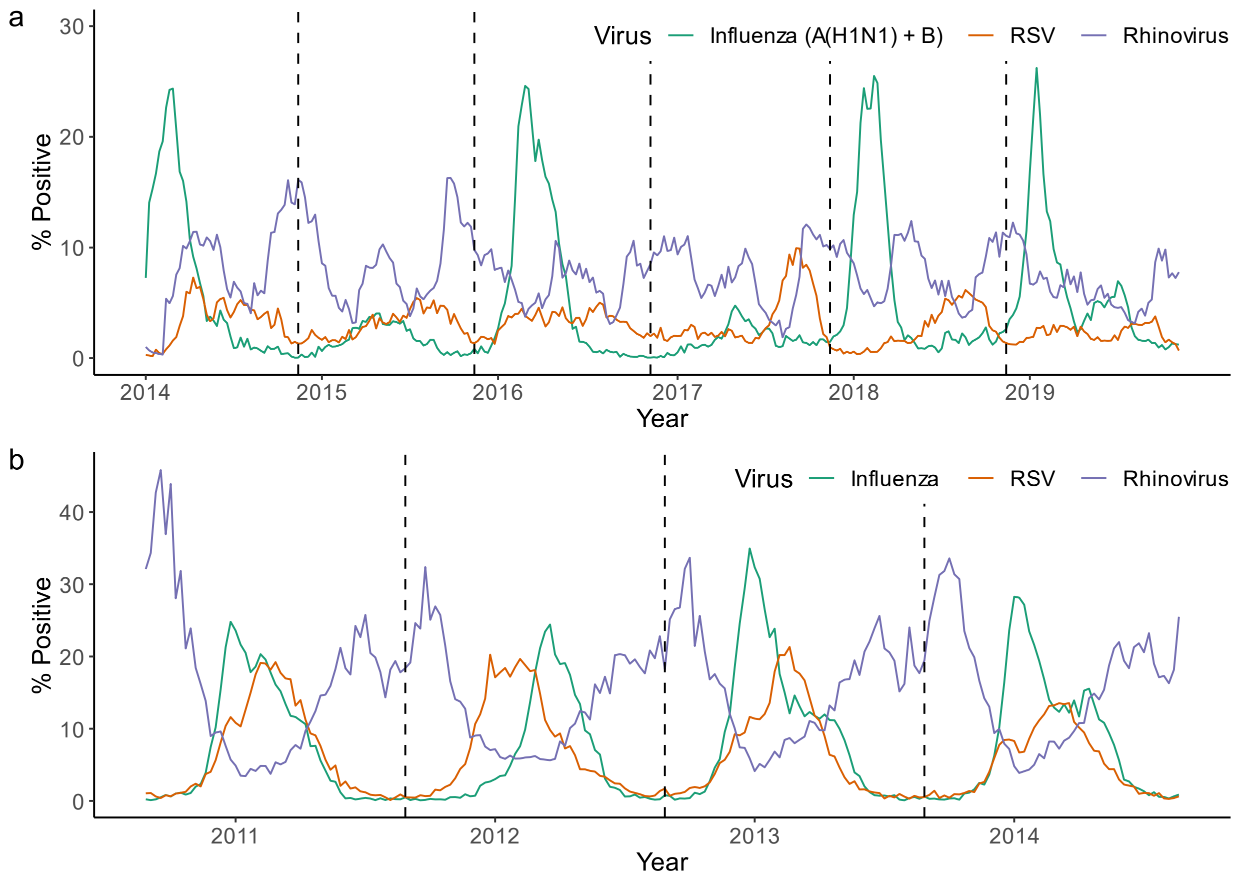


**Supplementary Figure 19. Percent of tests positive for influenza, RSV, and rhinovirus in Hong Kong (a) and Canada (b) throughout the study period.** Values for influenza and RSV are shown in green and orange, respectively, as in Fig. 1. Values for rhinovirus are shown in purple. In Hong Kong, influenza data only include cases of A(H1N1) and B influenza. Detailed information on how rhinovirus positivity was calculated for Hong Kong can be found in the Supplementary Text, under "Accounting for Circulation of Rhinovirus;" in Canada, the total number of tests conducted and positive for rhinovirus are reported directly.

**Supplementary Figure 20. Maximum likelihood estimates and 95% confidence intervals for shared parameters obtained from a variety of supplementary analyses.** Points show the maximum likelihood values, while error bars show the 95% confidence intervals, obtained using parametric bootstrapping with 500 synthetic datasets. Note that, instead of the duration parameters themselves ($\delta_1$ and $\delta_2$), the reciprocals of these parameters, indicating the duration in days of the interaction effect, are shown. Sensitivity analyses include: limiting the analysis to seasons where H3N2 circulation was minimal ("Low H3 Circulation"), assuming no interaction between influenza and RSV occurs ("No Interaction"), ignoring the potential impact of absolute humidity on transmissibility of both viruses ("Temperature Only"), modeling seasonal forcing using a sinusoidal wave rather than climate data ("Sinusoidal Forcing"), assuming that all members of the model population are fully susceptible to RSV at the beginning of each season ("$R_{20}$ + $R_{120}$ = 0"), and allowing rhinovirus incidence to modulate the transmissibility of influenza ("Rhino as Covariate"). Results from the main analysis are included for comparison ("Main"). All sensitivity analyses were conducted using the data from Hong Kong only.

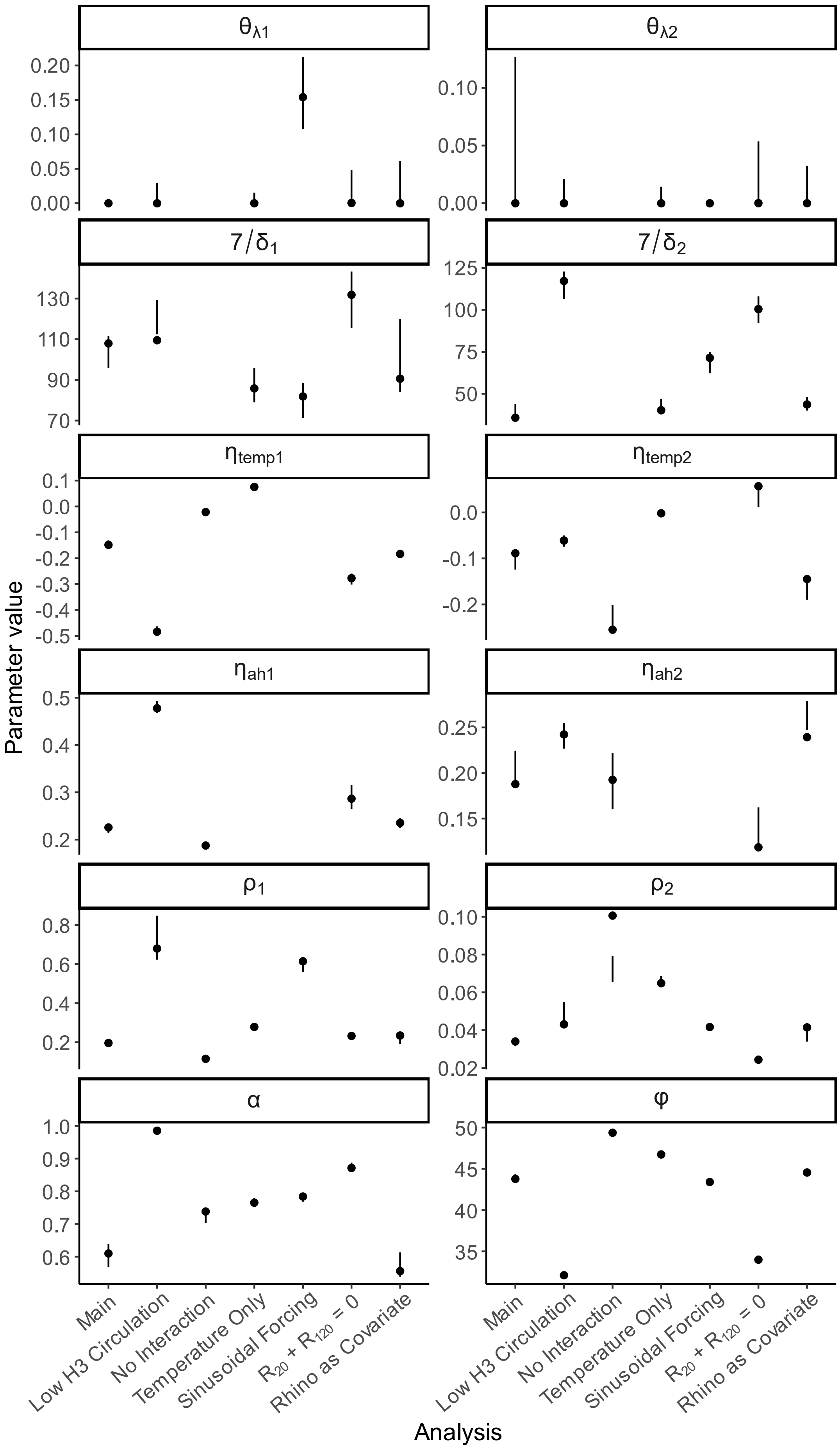

θλ1
θλ2
7/δ1
7/δ2
ηtemp1
ηtemp2
ηah1
ηah2
ρ1
ρ2
α
φ
Parameter value
Main
Low H3 Circulation
No Interaction
Temperature Only
Sinusoidal Forcing
R20 + R120 = 0
Rhino as Covariate
Analysis

### Supplementary Equations

**Supplementary Equation 1: Full model equations for the model of influenza and RSV cocirculation.**

$$\frac{dX_{SS}}{dt} = -(\lambda_1 + \lambda_2)X_{SS}$$

$$\frac{dX_{IS}}{dt} = \lambda_1 X_{SS} - (\gamma_1 + \theta_{\lambda 1}\lambda_2)X_{IS}$$

$$\frac{dX_{TS}}{dt} = \gamma_1 X_{IS} - (\delta_1 + \theta_{\lambda 1}\lambda_2)X_{TS}$$

$$\frac{dX_{RS}}{dt} = \delta_1 X_{TS} - \lambda_2 X_{RS}$$

$$\frac{dX_{SI}}{dt} = \lambda_2 X_{SS} - (\theta_{\lambda 2}\lambda_1 + \gamma_2)X_{SI}$$

$$\frac{dX_{II}}{dt} = \theta_{\lambda 1}\lambda_2 X_{IS} + \theta_{\lambda 2}\lambda_1 X_{SI} - (\gamma_1 + \gamma_2)X_{II}$$

$$\frac{dX_{TI}}{dt} = \theta_{\lambda 1}\lambda_2 X_{TS} + \gamma_1 X_{II} - (\delta_1 + \gamma_2)X_{TI}$$

$$\frac{dX_{RI}}{dt} = \lambda_2 X_{RS} + \delta_1 X_{TI} - \gamma_2 X_{RI}$$

$$\frac{dX_{ST}}{dt} = \gamma_2 X_{SI} - (\theta_{\lambda 2}\lambda_1 + \delta_2)X_{ST}$$

$$\frac{dX_{IT}}{dt} = \gamma_2 X_{II} + \theta_{\lambda 2}\lambda_1 X_{ST} - (\gamma_1 + \delta_2)X_{IT}$$

$$\frac{dX_{TT}}{dt} = \gamma_2 X_{TI} + \gamma_1 X_{IT} - (\delta_1 + \delta_2)X_{TT}$$

$$\frac{dX_{RT}}{dt} = \gamma_2 X_{RI} + \delta_1 X_{TT} - \delta_2 X_{RT}$$

$$\frac{dX_{SR}}{dt} = \delta_2 X_{ST} - \lambda_1 X_{SR}$$

$$\frac{dX_{IR}}{dt} = \delta_2 X_{IT} + \lambda_1 X_{SR} - \gamma_1 X_{IR}$$

$$\frac{dX_{TR}}{dt} = \delta_2 X_{TT} + \gamma_1 X_{IR} - \delta_1 X_{TR}$$

$$\frac{dX_{RR}}{dt} = \delta_2 X_{RT} + \delta_1 X_{TR}$$

**Supplementary Equation 2: Full model equations for the vaccination model.** Differences between these equations and those in Supplementary Equation (1) are shown in red.

$$\frac{dX_{SS}}{dt} = -(\lambda_1 + \lambda_2)X_{SS}$$

$$\frac{dX_{IS}}{dt} = \lambda_1 X_{SS} - (\gamma_1 + \theta_{\lambda 1}\lambda_2)X_{IS} + (1 - VE)\lambda_1(V_{SS1} + V_{SS2})$$

$$\frac{dX_{TS}}{dt} = \gamma_1 X_{IS} - (\delta_1 + \theta_{\lambda 1}\lambda_2)X_{TS}$$

$$\frac{dX_{RS}}{dt} = \delta_1 X_{TS} - \lambda_2 X_{RS} + \delta_{vacc}V_{RS}$$

$$\frac{dX_{SI}}{dt} = \lambda_2 X_{SS} - (\theta_{\lambda 2}\lambda_1 + \gamma_2)X_{SI}$$

$$\frac{dX_{II}}{dt} = \theta_{\lambda 1}\lambda_2 X_{IS} + \theta_{\lambda 2}\lambda_1 X_{SI} - (\gamma_1 + \gamma_2)X_{II} + (1 - VE)\theta_{\lambda 2}\lambda_1 V_{SI}$$

$$\frac{dX_{TI}}{dt} = \theta_{\lambda 1}\lambda_2 X_{TS} + \gamma_1 X_{II} - (\delta_1 + \gamma_2)X_{TI}$$

$$\frac{dX_{RI}}{dt} = \lambda_2 X_{RS} + \delta_1 X_{TI} - \gamma_2 X_{RI} + \theta_{\lambda_{vacc}}\lambda_2 V_{RS}$$

$$\frac{dX_{ST}}{dt} = \gamma_2 X_{SI} - (\theta_{\lambda 2}\lambda_1 + \delta_2)X_{ST}$$

$$\frac{dX_{IT}}{dt} = \gamma_2 X_{II} + \theta_{\lambda 2}\lambda_1 X_{ST} - (\gamma_1 + \delta_2)X_{IT} + (1 - VE)\theta_{\lambda 2}\lambda_1 V_{ST}$$

$$\frac{dX_{TT}}{dt} = \gamma_2 X_{TI} + \gamma_1 X_{IT} - (\delta_1 + \delta_2)X_{TT}$$

$$\frac{dX_{RT}}{dt} = \gamma_2 X_{RI} + \delta_1 X_{TT} - \delta_2 X_{RT}$$

$$\frac{dX_{SR}}{dt} = \delta_2 X_{ST} - \lambda_1 X_{SR}$$

$$\frac{dX_{IR}}{dt} = \delta_2 X_{IT} + \lambda_1 X_{SR} - \gamma_1 X_{IR} + (1 - VE)\lambda_1 V_{SR}$$

$$\frac{dX_{TR}}{dt} = \delta_2 X_{TT} + \gamma_1 X_{IR} - \delta_1 X_{TR}$$

$$\frac{dX_{RR}}{dt} = \delta_2 X_{RT} + \delta_1 X_{TR}$$

$$\frac{dV_{SS1}}{dt} = -\left(\delta_{vacc} + \theta_{\lambda_{vacc}}\lambda_2 + (1 - VE)\lambda_1\right)V_{SS1}$$

$$\frac{dV_{SS2}}{dt} = \delta_{vacc}V_{SS1} - \big((1 - VE)\lambda_1 + \lambda_2\big)V_{SS2}$$

$$\frac{dV_{SI}}{dt} = \theta_{\lambda_{vacc}}\lambda_2 V_{SS1} + \lambda_2 V_{SS2} - \big((1 - VE)\theta_{\lambda 2}\lambda_1 + \gamma_2\big)V_{SI}$$

$$\frac{dV_{ST}}{dt} = \gamma_2 V_{SI} - \big((1 - VE)\theta_{\lambda 2}\lambda_1 + \delta_2\big)V_{ST}$$

$$\frac{dV_{SR}}{dt} = \delta_2 V_{ST} - (1 - VE)\lambda_1 V_{SR}$$

$$\frac{dV_{RS}}{dt} = -\big(\delta_{vacc} + \theta_{\lambda_{vacc}}\lambda_2\big)V_{RS}$$

### Supplementary Text

1. Detailed model fitting methods

Model fitting to observed data was accomplished using a two-step process. First, we fit the model to each individual season of data separately, in order to obtain a reasonable first approximation for the values of the season-specific parameters. For each location and season, we fit the model 500 times, starting from 500 sets of initial parameter values drawn from broad but realistic ranges (Supplementary Table 2) using Latin hypercube sampling. This was done to ensure that the parameter space was adequately explored. In addition to the season-specific parameters, $\rho_1$ and $\rho_2$ were also fit in this step, as they partially represent reporting rates and fixing them could greatly compromise model fit; all other shared parameters were held constant.

Next, we fit the model to all seasons for a given location simultaneously, allowing season-specific parameters to vary by season while forcing shared parameters to take the same value in all seasons. Here, initial values for the season-specific parameters were selected from the best-fit ranges obtained in step one above. Initial values for shared parameters, including $\rho_1$ and $\rho_2$, were drawn from broad ranges (Supplementary Table 2). As in step one, we fit the model 500 times, starting from initial parameter value sets obtained using Latin hypercube sampling.

We then observed all fit parameter sets with values falling within the 95% confidence intervals, defined based on a chi-squared distribution with 54 (Hong Kong) or 40 (Canada) degrees of freedom (the total number of parameters fit), according to Wilks' theorem[1]. If only a single best-fit parameter set was identified, we assumed that convergence to the MLE had not yet occurred. In this case, we drew 500 new sets of initial parameter values from the ranges of parameter values found in the top 5% of fits and fit the model again. This process was repeated until more than one best-fit parameter set was obtained. At this point, a final round of model fitting was performed, with initial values drawn from the 95% confidence intervals for all parameters, as defined above. The best-fitting parameter set from this final run was taken to be the MLE. For the main analysis, this took a total of three runs in each location.

For the parametric bootstrap, each of the 500 synthetic outbreak sets was fit 10 times, using initial parameter values drawn from the best-fit parameter sets from the final round of model fitting described above. Only the best-fitting of the 10 runs for each synthetic outbreak set was used in calculating the 95% confidence intervals.

2. Sensitivity analyses

*Age-Structured Model*

In order to determine whether our homogenous mixing model is capable of correctly inferring interaction characteristics operating in a population with age structure, we conducted a sensitivity analysis in which we fit our model to synthetic data generated from an age-structured model. Specifically, we generated synthetic data using an age-structured model of influenza and RSV cocirculation. This model is broadly similar to the model used in the main text, with two primary differences: 1) transmission within and between age groups is dependent on observed age-specific contact rates between individuals, and 2) older age groups experience reduced susceptibility to RSV. The model population is divided into five age groups: less than one year old, 1-4 years old, 5-15 years old, 16-64 years old, and 65 years old and above.

To generate the synthetic, age-structured data, we set all model parameters to their MLEs, as obtained from fitting the homogenous mixing model to data on influenza and RSV circulation in Hong Kong (Table 1). Transmissibility values corresponding to the fit values of $Ri_1$ and $Ri_2$ were calculated using the next-generation matrix approach[2]. Values for the reduction in susceptibility to RSV by age were fixed to the same values as in Waterlow et al., such that susceptibility was reduced by 25% among those aged 1-4 years, and by 35% among those five years old and older. Mid-year population sizes for each age group in Hong Kong were obtained from the Census and Statistics Department of the Government of the Hong Kong Special Administrative Region[3]; as no data were available on the size of the population under the age of one, we assumed that this age group represented one fifth of the population aged 0 to 4. Age-specific contact rates for Hong Kong were obtained from Leung et al.[4]. Parameters for the reduction in susceptibility to RSV among older age groups were taken from Waterlow et al.[5]. We assigned the number of tests conducted for influenza and RSV each week for each age group by allocating the observed number of tests proportionally based on the observed rates of testing by age group in France[6], such that the total number of tests conducted each week was preserved. ILI rates per consultation were assumed to be the same in all age groups. This is a reasonable assumption: although we expect ILI consultations to be most prevalent among the youngest and oldest age groups, we also expect overall consultations to be highest in these groups.

We obtained counts of total influenza and RSV cases using the deterministic, age-structured model, then simulated from the stochastic observation model in order to generate weekly, age-specific counts of reported cases for all seasons. The resulting synthetic data are shown in Supplementary Fig. 18. We then summed the reported cases over all age groups to obtain the total number of reported cases in the population each week (Supplementary Fig. 17), and fit our homogenous mixing model to the summed data using the same model fitting approach as for our main analysis. The MLEs for all shared parameter values can be found in Supplementary Table 6. Overall, there is close agreement between the results of our main analysis and the age-structured sensitivity analysis described here, although the value of $\theta_{\lambda 1}$ is slightly overestimated (i.e., the strength of the interaction effect is slightly underestimated). Notably, this suggests that a failure to account for age structure may lead to a bias toward the null in the interaction strength parameters, at least in Hong Kong, suggesting that the interaction effect may be even stronger than what we have identified in the main text.

*Accounting for Circulation of H3N2*

To assess whether our decision not to account for H3N2 circulation in Hong Kong was likely to bias our estimates of the interaction parameters of interest, we conducted a sensitivity analysis in which we fit the model only to data from seasons where very little H3N2 was circulating (2017-18 and 2018-19; see Supplementary Fig. 2). Estimates of the interaction parameters obtained from these two seasons alone were very similar to those obtained when fitting to all available data, although the MLE of $d_2$ was 0.94, suggesting a similar duration of the interaction effect of RSV on influenza as of influenza on RSV (Supplementary Fig. 20). Overall, it appears that circulation of H3N2 has little effect on the estimated values of the interaction parameters.

*Sinusoidal Forcing*

As an alternative to explicitly incorporating observed temperature and absolute humidity into our modeled force of infection for influenza and RSV, we also fit a model where seasonal forcing was instead accomplished using a sinusoidal wave (see main text Equation (2)). Starting values for $b_1$ and $b_2$ were obtained by fitting a sine wave to the estimated extent of climate forcing at the MLE for each season, as obtained using the climate-forced model (Supplementary Fig. 8), and calculating the range of amplitudes.

Interestingly, we find that this model fit the data better than the climate-forced model used in our main analysis (Supplementary Table 5), although we continue to present the results from the climate forced model in the main text because of our interest in the fit values of the climate forcing parameters. The MLE values for the interaction parameters are similar to those obtained in the main analysis; however, the sinusoidal forcing model yields a slightly weaker interaction of influenza on RSV (0.15 vs. 0) with a shorter duration  (80 vs. 110 days), as well as a longer duration of the effect of RSV on influenza (70 vs. 36 days) than the climate-forced model (Supplementary Fig. 20).

*Accounting for Circulation of Rhinovirus*

We incorporated rhinovirus circulation (Supplementary Fig. 19) into our model by allowing rhinovirus incidence in a given week to modulate the force of infection of influenza during the same week.. Standardized rhinovirus incidence data were obtained by first multiplying the proportion of weekly tests positive for rhinovirus by the weekly rate of ILI. This was done to account for the fact that, because most symptomatic cases are not tested for rhinovirus, we expect the number of laboratory-confirmed cases to be a substantial underestimate of true rhinovirus circulation in the population. We then standardized the resulting values to have a mean of zero and a variance of one.

Unlike influenza and RSV, specific testing for rhinovirus was not conducted on all samples in Hong Kong. Rather, samples were tested to see whether they contained either rhinovirus or enterovirus. Then, some proportion of the positive samples were further tested to differentiate between rhinovirus and enterovirus.

We therefore estimated the weekly number of rhinovirus cases among those tested by multiplying the total number of cases positive for either rhinovirus or enterovirus by the proportion of samples each week that were further tested that contained rhinovirus specifically, and rounding to the nearest whole number. On average, about a third of samples positive for rhinovirus or enterovirus underwent further testing each week.

The weekly force of infection of influenza was then multiplied by:

$$\beta_{rhino}(t) = e^{-\beta_{rhino} inc_{rhino}(t)}$$

where $\beta_{rhino}$ represents the impact of rhinovirus on susceptibility to influenza and $inc_{rhino}(t)$ is the standardized incidence of rhinovirus during week *t*. Because rhinovirus is expected to inhibit influenza transmission[7,8], $\beta_{rhino}$ was constrained to be positive, such that increases in rhinovirus incidence would yield a decrease in the force of infection of influenza. Although this only allows us to account for a short-lived interaction between rhinovirus and influenza, it provides a simple, initial way of incorporating rhinovirus transmission into the model.

We found that the model accounting for rhinovirus transmission fit the data significantly better than did the model presented in the main text (log-likelihood = -9288.84). The MLEs of the interaction parameters were very similar for both models (Supplementary Fig. 20), suggesting that we are unlikely to be biasing our results by not accounting for rhinovirus transmission. The MLE for $\beta_{rhino}$ was 0.060 (95% CI 0.057-0.065), indicating that a one-unit increase in the standardized incidence of rhinovirus was associated with a 6% reduction in the force of infection of influenza. This analysis therefore supports the hypothesis that rhinovirus is able to suppress susceptibility to influenza, although, as we note in the main text, further research using models designed to explicitly account for rhinovirus circulation will be instrumental in understanding this relationship more completely.

3. Interpretation of the composite reporting rate and scaling parameters, $\rho_1$ and $\rho_2$

We want $\rho_1$ and $\rho_2$ to represent the probability that an individual in the model population is infected with influenza or RSV, respectively, given that they have ILI. Here, we infer the form of $\rho_1$ only, although the form of $\rho_2$ can be inferred similarly. According to Bayes' Rule:

$$p(influenza|ILI) = \frac{p(ILI|influenza)p(influenza)}{p(ILI)}$$

The quantity $p(influenza)$ represents the probability that an individual in the population is infected with influenza, and is the quantity output by our model ($H_1(t)$). Meanwhile, the observed ILI data from Hong Kong and Canada ($ILI(t)$) are reported as the proportion of all consultations that are due to ILI, such that:

$$ILI(t) = \frac{p(ILI)N}{C(t)}$$

where $N$ represents the total population size, and $C(t)$ represents the number of all-cause consultations during week *t*. Combining these equations, we get:

$$p(ILI) = \frac{p(ILI|influenza)}{p(consultation)} \frac{H_1(t)}{ILI(t)}$$

Hence, $\rho_1$ is of the form:

$$\rho_1 = \frac{p(ILI|influenza)}{p(consultation)}$$

where $p(ILI|influenza)$ represents the probability that an influenza case is diagnosed with ILI, and $p(consultation)$ represents the probability of attending a public out-patient clinic for any reason. The former term accounts for both the probability that an influenza case develops symptoms consistent with ILI, as well as the probability that an ILI case results in a consultation and diagnosis. The latter term, meanwhile, allows for conversion between ILI rates per consultation, as reported in the data, and ILI rates per population, as output by the model[9].

#### Supplementary Tables

**Supplementary Table 1.** Observed epidemic characteristics.

| Metric | Virus | Median Value (Range) | |
|---|---|---|---|
| | | Hong Kong | Canada |
| % of All Tests Positive | Influenza | 7.1% (1.7%-8.2%) | 13.3% (9.1%-16.7%) |
| | RSV | 3.2% (2.2%-3.5%) | 9.6% (7.1%-10.0%) |
| Peak Week | Influenza | 9 (3-18) | 52.5 (52-12) |
| | RSV | 32.5 (15-38) | 8.5 (52-12) |
| Peak Week Difference* | Influenza | -20 (-35 - -6) | -8.5 (-11 - 12) |
| | RSV | NA | NA |
| Duration** | Influenza | 16 (10-26) | 11.5 (11-13) |
| | RSV | 29 (24-32) | 14.5 (14-16) |

*Peak Week of influenza minus Peak Week of RSV; negative values indicate that influenza peaks before RSV
**The number of weeks containing at least 75% of the total number of laboratory-confirmed cases occurring in a season; weeks are not necessarily consecutive

**Supplementary Table 2.** Range of initial values used for model fitting for all parameters.

| Parameter | Initial Value Range | Season-Specific? |
|---|---|---|
| $\theta_{\lambda 1}$ | 0 – 1.0 | No |
| $\theta_{\lambda 2}$ | 0 – 1.0 | No |
| $\delta_1$ | 7 / 60 – 7 | No |
| $d_2$ | 0 – 10.0 | No |
| $\rho_1$ | 0 – 1.0 | No |
| $\rho_2$ | 0 – 1.0 | No |
| $\eta_{temp1}$ | -0.5 – 0.5 | No |
| $\eta_{AH1}$ | -0.5 – 0.5 | No |
| $\eta_{temp2}$ | -0.5 – 0.5 | No |
| $\eta_{AH2}$ | -0.5 – 0.5 | No |
| $b_1$ | 0.05 - 0.20 | No |
| $b_2$ | 0.05 - 0.20 | No |
| $\varphi_1$ | 0 – 52.25 | No |
| $\varphi_2$ | 0 – 52.25 | No |
| $\alpha$ | 0 – 0.5 | No |
| $\varphi$ | 0 – 52.25 | No |
| $Ri_1$ | 1.0 – 3.0 | Yes |
| $Ri_2$ | 1.0 – 3.0 | Yes |
| $I_{10}$ | 0 – 0.001 | Yes |
| $I_{20}$ | 0 – 0.001 | Yes |
| $R_{10}$ | 0 – 0.3 | Yes |
| $R_{20}$ | 0 – 0.3 | Yes |
| $R_{120}$ | 0 – 0.3 | Yes |

**Supplementary Table 3.** Nash-Sutcliffe coefficient of efficiency ($R^2$) comparing the fit of the model presented in the main text to fits of a sine wave through all data for each location. Positive values indicate that the mechanistic model outperforms the sine wave, whereas negative values indicate that the sine wave demonstrated better-quality fit.

| Virus | Coefficient of efficiency ($R^2$) | |
|---|---|---|
| | Hong Kong | Canada |
| Influenza | 0.90 | 0.91 |
| RSV | 0.53 | -0.04 |

**Supplementary Table 4.** Percent of 100 simulations at the maximum likelihood estimate that accurately reproduce outbreak dynamics observed in the case count data. Mean values and the full range across seasons are shown. Peak timing refers to the week at which the number of observed cases was maximal, peak intensity is the number of cases observed during the peak week, and attack rate is the total number of cases observed throughout a given season. Peak timing is considered to be accurate if the predicted peak is within 2 weeks of the observed peak; peak intensity and attack rate are accurate if the predicted values are within 25% of the observed value.

| Outbreak Metric | Virus | % Accurate (Seasonal Range) | |
|---|---|---|---|
| | | Hong Kong | Canada |
| Peak Timing | Influenza | 85.8% (39-100) | 94.8% (80-100) |
| | RSV | 25.5% (0-97) | 70.0% (0-100) |
| Peak Intensity | Influenza | 77.0% (0-100) | 65.8% (22-85) |
| | RSV | 83.5% (12-100) | 63.5% (2-100) |
| Attack Rate | Influenza | 99.7% (98-100) | 91.3% (73-100) |
| | RSV | 90.5% (43-100) | 100% (100-100) |

**Supplementary Table 5.** Log-likelihood values and AICs of the best-fit model parameter sets for the model used in the main text ("Main") and for several sensitivity analyses. Models were compared using one-sided likelihood-ratio tests, where the degrees of freedom (df) were the difference in the number of included parameters as compared to the "Main" model. A significant difference in model fit compared to the main model is indicated with an "*" next to the log-likelihood; all differences were significant with $p < 1\times10^{-10}$.

| Model | Log-likelihood | df | AIC |
|---|---|---|---|
| Main | -9681.43 | NA | 19470.85 |
| Temperature Only | -10675.50* | 2 | 21454.99 |
| Sinusoidal Forcing | -8853.06† | 0 | 17814.13 |
| $R_{20} = R_{120} = 0$ | -10628.02* | 12 | 21340.04 |
| No Interaction | -12213.04* | 4 | 24526.07 |

†Because the sinusoidal forcing model is not nested in the main model, we cannot assess significance using a likelihood ratio test. However, the difference in the AICs suggest that this model fits the data meaningfully better than the main model.

**Supplementary Table 6.** Maximum likelihood estimates from the age-structured sensitivity analysis, compared to true values (from Table 1), for all shared model parameters

| Parameter | MLE | True Value |
|---|---|---|
| $\theta_{\lambda1}$ | 0.044 | $1.2\times10^{-9}$ |
| $\theta_{\lambda2}$ | 0.074 | $6.2\times10^{-6}$ |
| $\delta_1$ | 0.060 | 0.065 |
| $d_2$ | 2.80 | 3.02 |
| $\rho_1$ | 0.166 | 0.20 |
| $\rho_2$ | 0.035 | 0.034 |
| $\eta_{temp1}$ | -0.13 | -0.15 |
| $\eta_{AH1}$ | 0.23 | 0.23 |
| $\eta_{temp2}$ | -0.11 | -0.09 |
| $\eta_{AH2}$ | 0.22 | 0.19 |
| $\alpha$ | 0.58 | 0.61 |
| $\varphi$ | 42.9 | 43.8 |
